# Optimising covariate allocation at design stage using Fisher Information Matrix for Non-Linear Mixed Effects Models in pharmacometrics

**DOI:** 10.1101/2025.01.31.25321452

**Authors:** Lucie Fayette, Karl Brendel, France Mentré

## Abstract

This work focuses on designing experiments for pharmacometrics studies analysed by Non-Linear Mixed Effects Models (NLMEM) including covariates to describe inter-individual variability. Co-variate effects may help identify patient subpopulations at risk of sub-therapeutic or toxic responses, for instance when hepatic or renal impairment reduces drug elimination, increasing safety risks. Before collecting and modelling new clinical trial data, choosing an appropriate design is crucial, particularly to ensure sufficient information to estimate covariate effects and their uncertainty. Assuming a known NLMEM with covariate effects and a joint distribution for covariates in the target population from previous clinical studies, we propose to optimise the allocation of covariates among the subjects to be included in a new trial. It aims achieving better overall parameter estimations and therefore increase power of statistical tests on covariate effects to detect the clinical relevance or non-relevance of relationships. We used the Fisher Information Matrix and developed a fast and deterministic computation method, leveraging Gaussian quadrature and copula modelling. We suggested dividing the domain of continuous covariates into clinically meaningful intervals and optimised their proportions, along with the proportion of each category for the discrete covariates. The optimisation problem was formulated as a convex problem subject to linear constraints, allowing resolution using Projected Gradient Descent algorithm. Different scenarios for a pharmacokinetics model including either biological measurements of renal or hepatic function as covariate were explored. We showed that covariate optimisation reduces the number of subjects needed to achieve desired power in covariate tests for relevance or non-relevance.

**Highlights:**

- Fisher Information Matrix in nonlinear mixed effects models (NLMEM) with covariates
- New computation method of Information Matrix using copula and Gaussian quadrature
- Optimisation of covariate allocation using partitioning and projected gradient
- Reduced sample size needed to detect clinically relevant covariate in a PK example

## 1. Introduction

In pharmacometrics longitudinal data collected during clinical trials are analysed through Non-Linear Mixed Effect Models (NLMEM). Notably, NLMEM allow to quantify the relationships between covariates and parameters (Mould and Upton, 2012). Indeed, covariates are often included in pharmacometrics models to describe predictable between inter-individual variability (IIV), thereby enhancing model fit and/or model-based predictions (Sanghavi et al., 2024). Covariates are individual-specific characteristics known at baseline, which may be either discrete (e.g., sex, ethnicity) or continuous (e.g., body weight, creatinine clearance). In addition, covariate effects may help identify patient subpopulations at risk of sub-therapeutic or toxic responses, thus supporting the individualization of drug therapy (Joerger, 2012). Especially, in pharmacokinetics (PK) covariates can have effects on drug exposure requiring dose adjustment whose clinical relevance must be proven (FDA, 2022). This is particularly true for renal or hepatic impairment which can reduce the elimination of drugs and lead to increased safety risks (FDA, 2024, 2003; EMA, 2016, 2005). These dysfunctions can be quantified using one or more correlated biological covariates. When clinical studies enrolled a diverse patient population covering the full range of covariate values, and when sufficient PK data are available to evaluate the impact of organ impairment, population PK analyses from phase 2 and/or phase 3 trials may be sufficient to estimate the effect of renal function (RF) and/or hepatic function (HF) on drug exposure and its uncertainty within the studied population (FDA, 2024). Otherwise, dedicated additional studies should be conducted. Regulatory guidance recommends the use of confidence intervals to interpret change of exposure as ratios between groups (FDA, 2024, 2003; EMA, 2016, 2005), thereby supporting a quantitative assessment of potential PK alterations.

Our objective is therefore to support the design of future clinical trials in pharmacometrics by computing optimal covariate distributions that ensure sufficient statistical power to detect clinically relevant or non-relevant covariate relationships. In particular, we aim to determine the required number of subjects within subpopulations of interest, and thus avoid conducting specific trials to assess the effect of specific impairments on PK.

Before collecting and modelling the data, a key step is indeed to choose an appropriate design. In NLMEM, a population design refers to the number of subjects and their allocation to elementary designs, an elementary design being a combination of values for the design variables, such as number of samples, sampling times, dosage regimen, etc. The aim of the design phase is thus to improve the structure of a study to maximize the informativeness of the data that would be collected. This approach is particularly valuable when dealing with sparse sampling (e.g., paediatrics, rare diseases), ethical or logistical constraints or high IIV. Usually, covariates are not considered at this step, and although it is commonly recommended to include subjects spanning the full range of covariate values (FDA, 2022; Sanghavi et al., 2024), there is currently no method for quantifying the influence of their distribution within the range. In its Population PK Guidance for Industry 2022 (FDA, 2022), the FDA emphasises that “Simulations and optimal design methods can maximize the utility of population PK data collection and analyses” and stresses in particular that these methods can aim to estimate “major covariate effects of interest” with greater precision. Nevertheless, the method proposed for power assessment and sample size computation in this guidance is clinical trial simulations (CTS), although it is computationally expensive and less exhaustive than optimal design strategies.

The traditional theory of optimal experimental design, initially developed for non-linear models (Atkinson et al., 2007), have been imported to the field of NLMEM and pharmacometrics (Mentre et al., 1997; Mentré et al., 2013; Strömberg et al., 2016) and relies on Fisher Information Matrix (FIM) computation. In the NLMEM framework, the FIM does not have a closed-form solution, requiring the computation of two intractable integrals: the integration of the derivatives of the log-likelihood over the random effects, and the evaluation of their expectation with respect to the observations. For continuous responses, a first-order linearisation of the model around 0, the mean of the random effects, results in a Gaussian approximation for which an analytical formula can be computed (Mentre et al., 1997; Bazzoli et al., 2009). Otherwise, the expectation over the observations can be approximated through a Monte Carlo procedure coupled with a linearisation of the model around the empirical Bayes estimate of the random effects, to bypass the integration over the random effects (Retout and Mentré, 2003). For design purpose with continuous responses, the linear approximation has been shown to be appropriate (Strömberg et al., 2016). If the response is discrete or if a linear approximation is not appropriate, other methods have been proposed, such as Markov Chain Monte Carlo (MCMC) to integrate over the random effects (Riviere et al., 2016), or Adaptive Gaussian Quadrature (for continuous (Nguyen and Mentré, 2014) or discrete (Ueckert and Mentré, 2017) outcomes), both of which combine Monte Carlo methods to evaluate the expectation over the observations. In those works, covariates were generally either absent or fixed, except in (Retout and Mentré, 2003), where the integral over the covariate distribution was computed using a Monte Carlo procedure. Indeed, when covariates are treated as random variables, the FIM becomes an expectation over both the observations and the covariates. As a result, an additional integral must be computed with respect to the distribution of the covariate vector. The linearisation approach has been implemented in various software tools (Nyberg et al., 2015). Thus, before the start of a new study, given a NLMEM and parameters value, one can compute the expected standard error (SE) of the model parameters and deduce whether enough information will be collected to meet the objectives of the study. Thereafter, design optimisation consists in finding the design of experiments that provides the most informative data for estimating model parameters while respecting some design constraints, such as number of subjects, number of sampling times or samplings times windows.

Nonetheless, accounting for covariates and their distribution during the design phase enables the quantification of how both the variations in covariates and the design influence the confidence intervals of covariate effects and the statistical power of the associated tests. It also enables assessment of the impact of covariate distributions on parameter identifiability and the determination of the minimum sample size required to achieve a desired statistical power. However, this aspect has not been adequately addressed in the literature, and existing software tools handle only known and fixed covariate values, rather than full covariate distributions. Previous works have looked at how to account for covariates when allocating patients to different arms, and how an unbalanced design could improve efficiency and ethics, including model-based approaches (see for instance (Rosenberger and Sverdlov, 2008) for a review). Nevertheless, these approaches do not use the longitudinal data framework nor NLMEM. In addition, they aim to optimise the design sequentially and seek, for each new patient entering the trial, to allocate them to the best possible arm in terms of efficacy and ethical considerations, while maintaining randomisation. Consequently, it does not correspond to our setting aiming to optimise the design prior to setting up the trial. In (Bogacka et al., 2017), a new method based of the FIM for optimising design using NLMEM where covariates are design variables is proposed. However, the proposed methods require the FIM to be evaluated for a known set of covariate values, without taking into account the distribution of these covariates, so the combinatorial approach can result in a very large number of elementary designs to be evaluated, and the optimisation result is discrete, making it difficult to manage continuous covariates.

In a previous work, we explored methods for handling covariates distribution in NLMEM with FIM computed by linearisation (Fayette et al., 2025). The integral with respect to covariate distribution was approximated using Monte Carlo integration, and we proposed to leverage either an historical covariate dataset, a known covariate distribution or a copula. This approach enables simultaneous handling of both discrete and continuous covariates, while accounting for their correlations. The accuracy in predicting uncertainty and power of significance and relevance tests was assessed in a simulation study. This approach was also used to optimize the sampling scheme in an in silico study comparing imputation methods for missing data (Duflot et al., 2025).

In the present work, we developed a new method aimed at reducing the computational cost of the integration with respect to covariate distribution in the linearised FIM. Indeed, with optimisation purposes, the required number of FIM evaluation increases and run time becomes a challenge. Therefore, avoiding Monte Carlo simulations is a lever to reduce this cost. As a consequence, we introduce a Gaussian quadrature (Golub and Welsch, 1969) (GQ) based approach, thus faster and deterministic. We also developed a workflow to determine the number of quadrature nodes required to ensure accurate FIM computation. We have coupled this approach with the use of copula to describe the distribution of covariates, as they are very flexible and, if already available, can be used without individual data.

Regarding optimisation, a hurdle to be overcome is that optimising the distribution of continuous covariates is not trivial as it is unrealistic to recommend including a given number of patients matching an exact value of the covariate. Therefore, we propose to segment the continuous covariates into different meaningful intervals and to optimise the proportion of each of them among the trial population. The combination of possible continuous intervals and discrete categories defines a finite number of subgroups describing the entire population. This has the combined advantage to remains realistic in practice while defining discrete parameters to be optimised. Thus, the covariate distribution in the population is here defined as a mixture of subgroups distributions, and we propose a method to construct an enrolment strategy based on the weighting of these subgroups. The FIM is computed over a finite set combining elementary designs and covariate distributions and each combination is optimally weighted, given the constraints, using the Projected Gradient Descent (PGD) algorithm (Calamai and Moré, 1987). Once the covariate distribution has been optimised, the sample size can be estimated to achieve the desired level of precision, as well as the number of subjects needed (NSN) to attain the desired power for relevance tests on covariate effects.

Section 2 details the notations and methods developed in this work. In Section 3, we present an example based on a one-compartment PK model, exploring various scenarios to demonstrate the benefits of optimizing covariate distributions for detecting the relevance or non-relevance of covariate effects in the context of renal or hepatic impairment. Since prior data or copulas are not always available at the design stage, we used publicly available data from the National Health and Nutrition Examination Surveys (NHANES) database (Centers for Disease Control and Prevention (CDC).National Center for Health Statistics (NCHS), 2009-2020) to fit copula as prior covariate distributions. Otherwise, data available from previous studies may also be used. The results are presented in Section 4 and discussed in Section 5.

## 2. Methods

In this section we detail notations and methods on study design and NLMEM. We then describe the FIM integration using copula and Gauss-Legendre Quadrature (GLQ). Next, we present the optimisation of covariate distributions, with a particular focus on the segmentation of continuous covariates for optimisation purposes and how it is handle in the FIM computation. Finally, we in-troduce the computation of ratios used to assess the clinical relevance or non-relevance of covariate relationships.

### 2.1. Notations on study designs and NLMEM

In the following, a population design is denoted Ξ = {*N*, (*ξ*_1_, …, *ξ*_*N*_)}, where *N* is the number of subjects and the *ξ*_*i*_ are their elementary designs.

The observation vector *y*_*i*_ for the *i*^*th*^ subject is modelled as given in equation (1) where *f* denotes the structural non-linear model and *θ*_*i*_ = *u* (*µ, β, Z*_*i*_, *η*_*i*_), the *p*-vector of individual parameters. The latter are defined as a function *u*, depending on fixed effects, composed of the typical values denoted *µ* and of the covariate effects *β*, and on individual random effects *η*_*i*_, where *η*_*i*_ *∼* 𝒩 (0, Ω). Individual parameters also depend on the vector of possibly transformed individual covariates *Z*_*i*_.

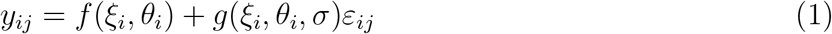

The residual error is modelled by the function *g*, depending on *ξ*_*i*_, *θ*_*i*_ and some parameters *σ*, and by the random variable *ε*_*i*_, with 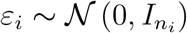.

We denote *Z*_*i*_ = (*Z*_*Dis,i*_, *Z*_*Cont,i*_) a covariate vector when considered as a random vector, and *z*_*i*_ = (*z*_*Dis,i*_, *z*_*Cont,i*_) as a realisation of this random vector. The lower._*Dis*_ refers to the discrete subvector of the covariate vector, while._*Cont*_ refers to the continuous sub-vector. In pharmacometrics modelling, parameters are generally log-normally distributed with additive covariate relationships on the log scale, i.e. log *θ*_*i*_ = log *µ* + *βz*_*i*_ + *η*_*i*_.

The *P* -vector of population parameters is denoted Ψ = {*µ, β, λ*}, where *λ* contains the variance parameters (elements of Ω and of the residual error model *σ*).

### 2.2. Elementary and population FIM

Given an elementary design *ξ*_*i*_, a population parameters vector Ψ, and a covariate realisation *z*_*i*_ the expected elementary FIM, *M*_*F*_ (*ξ*_*i*_, Ψ, *z*_*i*_), is defined as the covariance matrix of the Fisher score (equation (2)) where *l*(*y*_*i*_; Ψ, *z*_*i*_, *ξ*_*i*_) is the likelihood of the vector of observations *y*_*i*_ for the population parameters Ψ, given the individual vector of covariates *z*_*i*_ and the elementary design *ξ*_*i*_ (equation (3)).

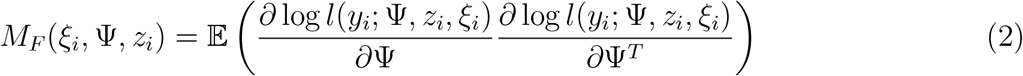

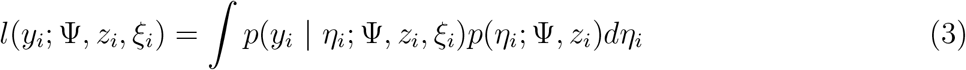

Knowing the distribution *p*_*Z*_ for the covariate vector, the expected FIM can be expressed as an expectation over the covariate vector as given in equation (4).

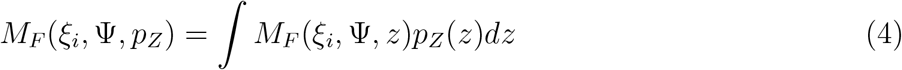

Assuming that the covariate distribution *p*_*Z*_ is the same for all the elementary design, the expected population FIM is the sum of the elementary FIM (equation (5)).

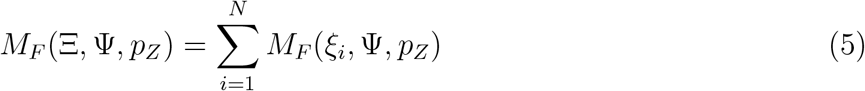

In the following, for sake of simplicity we consider that the *N* subjects have the same elementary design *ξ*, thus the expected population FIM is *M*_*F*_ (Ξ, Ψ, *p*_*Z*_) = *NM*_*F*_ (*ξ*, Ψ, *p*_*Z*_).

In the case with only *D* discrete covariates, each with *Q*_*δ*_, *δ* = 1, …, *D* categories. The covariate vector follows a discrete distribution with 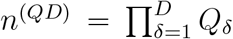 possible values, denoted *z*_*Dis,d*_, *d* = 1, …, *n*^(*QD*)^, each having the probability *p*_*Dis,d*_, *d* = 1, …, *n*^(*QD*)^. Therefore, the elementary FIM for *ξ* writes as a weighted sum as given in equation (6).

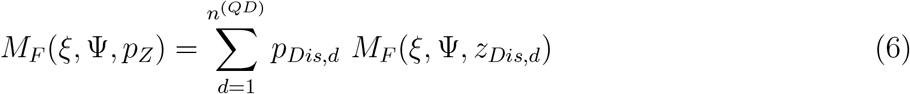

In case there are both non-independent discrete and continuous covariates, for each of its possible value *z*_*Dis,d*_, we denote 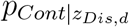 the distribution of the continuous sub-vector conditionally to *z*_*Dis,d*_. The elementary FIM for a given *ξ* is given in equation (7).

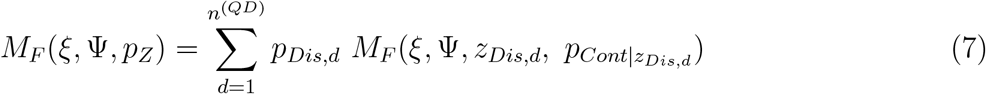

### 2.3. FIM integration using copula and Gauss-Legendre Quadrature

To compute the elementary FIM given in equation (7), the *n*^(*QD*)^ matrices 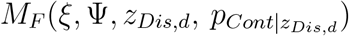 must be evaluated. For the sake of simplicity, we omit the index, *d* in the remainder of this paragraph. The first aspect of our method is to characterise the covariate distribution through copula modelling to account for correlations in FIM computation as already proposed (Fayette et al., 2025). Indeed, to perform GLQ integration, an explicit formula for the joint density of the covariates is required. We opted to use copulas rather than assuming a pre-specified multivariate distribution (e.g., multivariate Gaussian), due to their greater flexibility in capturing complex dependency structures.

It is well known that given a random vector (*Z*_1_, *Z*_2_, …, *Z*_*C*_) which follows the distribution 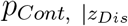 and has continuous marginals *F*_1_, …, *F*_*C*_, the random vector (*U*_1_, *U*_2_, …, *U*_*C*_) = (*F*_1_(*Z*_1_), *F*_2_(*Z*_2_), …, *F*_*C*_(*Z*_*C*_)) has marginals uniformly distributed on [0; 1]. By definition, the copula of (*Z*_1_, *Z*_2_, …, *Z*_*C*_) is the cumulative distribution function (cdf) of (*U*_1_, *U*_2_, …, *U*_*C*_). The Rosenblatt transform (Rosenblatt, 1952) denoted 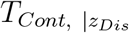 is defined in equation (8) and its inverse in equation (9).

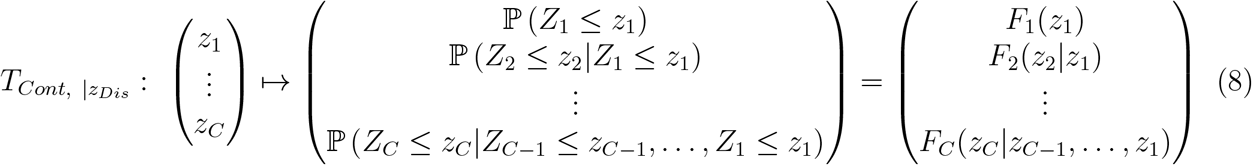

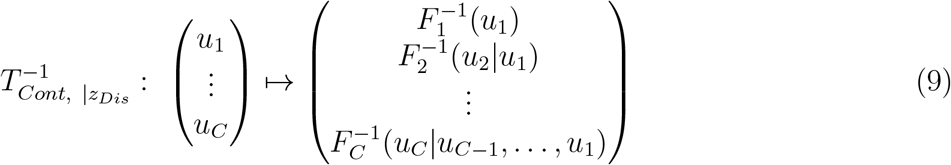

Therefore, applying the inverse Rosenblatt transform to a vector of random uniform variables re-turn a vector that has the same distribution as (*Z*_1_, *Z*_2_, …, *Z*_*C*_). Consequently, 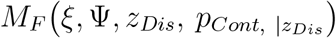 can be expressed using 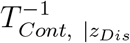 as given in equation (10).

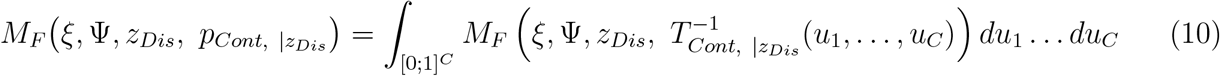

The second aspect is to approximate this integral through GQ (Golub and Welsch, 1969). The latter consists in approximating an integral by a weighted sum of the function to be integrated, evaluated at a number *n*^(*Q*)^ of given points in the integration domain, called quadrature nodes. The advantage of GQ integration over the Monte Carlo is that, since the nodes are chosen to represent the distribution harmoniously, equivalent accuracy is achieved with fewer GQ nodes than with random draws for Monte Carlo (James, 1980).

In our case the integration domain for each of the *C* integrals is [0; 1]. Therefore, we apply *C* GLQ with a substitution, as GLQ allows to integrate over the interval [-1, 1]. Denoting *v*_*q*_ and *w*_*q*_ the *q*^*th*^ node and its weight in GLQ, the substitutions are: 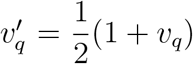 and 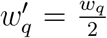. Finally, 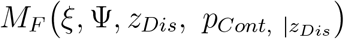 can be expressed as given in equation (11).

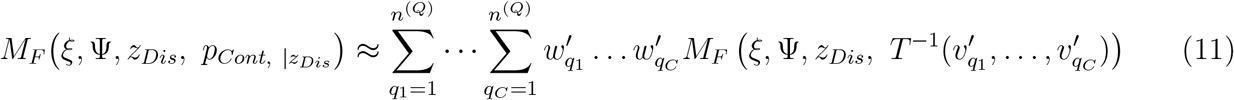

### 2.4. Optimisation of covariate distributions

We assume here that initial distributions are known for the covariates, and our aim is to optimally allocate subjects within the study accordingly. In this subsection, we describe first the segmentation of continuous covariates for optimisation, then the optimisation criteria, the proposed optimisation algorithm and the derivation of *NSN*.

#### 2.4.1. Segmentation of continuous covariates

In order to optimise the allocation for continuous covariates, we suggest segmenting the continuous covariates into different intervals, chosen for their clinical meaning. We then distribute the allocation between the intervals of continuous covariates and between the categories of discrete covariates. The advantage is that it preserves the continuous nature of the covariates in the FIM calculus, while providing a discrete segmentation. Such segmentation is advantageous for reducing dimensionality during optimisation and for guiding subject inclusion based on the optimisation results.

We consider a general framework, with respectively *D* discrete and *C* continuous covariates.

The domain of each continuous covariate, indexed by *c*, is segmented in 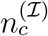 intervals denoted 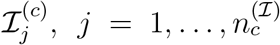. Combining those continuous intervals leads to 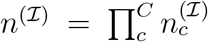 possible intervals for *Z*_*Cont*_. Finally, there are *L* = *n*^(*QD*)^ *× n*^(ℐ)^ possible distributions for the covariate vector *Z*. Conditionally to *Z*_*Dis*_ = *z*_*Dis,d*_, the distribution of 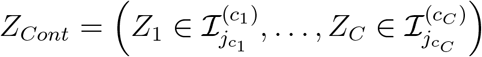 is denoted 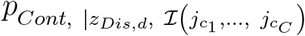. The proportion of subjects within the trial population and for which *Z*_*Dis,i*_ = *z*_*Dis,d*_ and 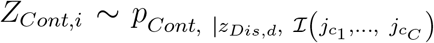 is denoted: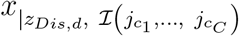. This segmentation leads to the elementary FIM formula given in equation (12), which now depends on ***x***.

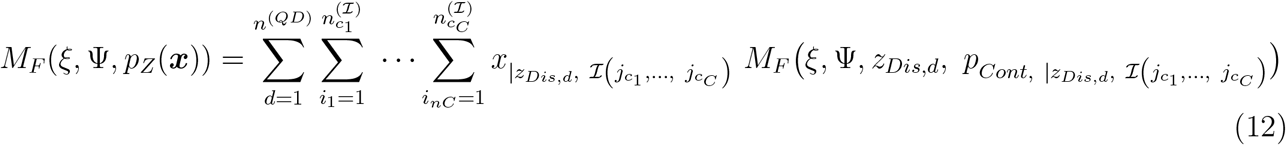

We denote ***x*** the vector of size *L* containing the 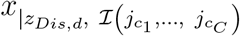 and for a better clarity in the notations, we denote *x*_*l*_ the *l*^*th*^ element of ***x*** and *p*_*Z,l*_ the associated distribution for *Z*. The overall mixture distribution for *Z* is denoted *p*_*Z*_(***x***). These notations result in the FIM formula given in equation (13). Finally, the multivariate distribution of covariates within the trial is described as the mixture of *L* multivariate distributions, and it is these mixture weights that we are seeking to optimise.

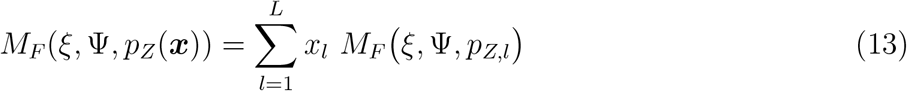

Each of the *L* matrices that appears in equation (13) can be computed using the GLQ approach presented above.

#### 2.4.2. Optimisation criterion

In order to compare the FIM between different designs, a criteria mapping the matrix to a scalar needs to be defined. Among various optimality criterion used in theory of optimal design (see for instance (Atkinson et al., 2007)), the D-criterion is a popular one and is wieldy used in the field of pharmacometrics. It uses the determinant of the FIM, as given in equation (14) and maximizing it ensures that the parameter estimates are as precise as possible. Indeed, a larger determinant of the FIM corresponds to a smaller volume of the confidence ellipsoid for the parameter estimates, indicating higher precision.

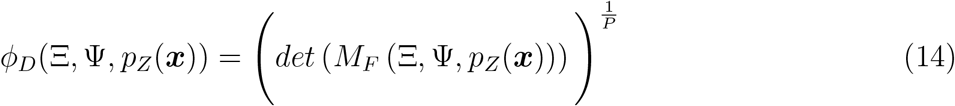

The comparison of covariate repartition ***x*** against a reference ***x***_*ref*_ for the D-criterion is done by calculating the D-efficiency *E*_*D*_ (equation (15)).

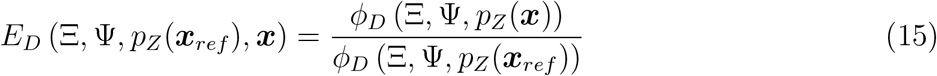

#### 2.4.3. Optimising covariate proportions using Projected Gradient Descent

Our aim is to maximize the D-criterion, for a fixed design Ξ by distributing the various covariate distributions weighted by the *x*_*l*_, and given some constraints on these proportions.

##### Linear constraints

First, as ***x*** represents a vector of proportions, it should sum to 1 and each *x*_*l*_ has to be in [0; 1]. Additional constraints can be set on the *x*_*l*_, for instance to make sure that a given interval for a continuous covariate does not represent more than a desired proportion and not less than another proportion. Therefore there are both equality and inequality constraints, and the optimisation problem is given in equation (16). We denote *Q* the constraint space.

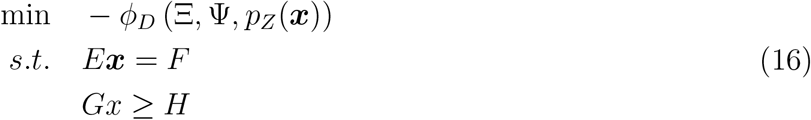

##### Projected Gradient Descent

As the D-criterion is a convex function, the PGD (Calamai and Moré, 1987) can be used to solve the problem given in equation (16). Starting from an initial point *x*_0_ ∈ *Q*, PGD iterates the equation given in (17), where *α*_*k*_ ∈ [0; ∞] is the gradient step-size at the *k*^*th*^ iteration, ∇ *ϕ*_*D*_ the gradient of *ϕ*_*D*_ with respect to ***x*** and 𝒫 _𝒬_ is the projection on the constraint space. This projection is also the solution of a minimization problem, given in equation (18).

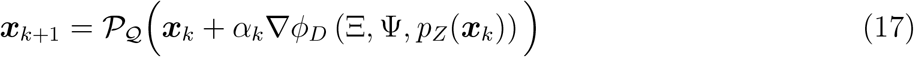

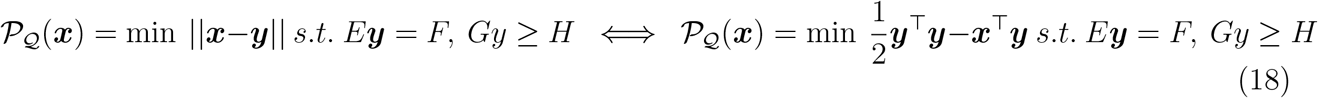

PGD iterates until a stopping condition is met. The detailed computation for ∇*ϕ*_*D*_ (Ξ, Ψ, *p*_*Z*_, ***x***_*k*_) when all the subjects have the same elementary design *ξ* is given in (Fayette et al., 2023) and the result is recall in equation (19), where tr denotes the trace operator.

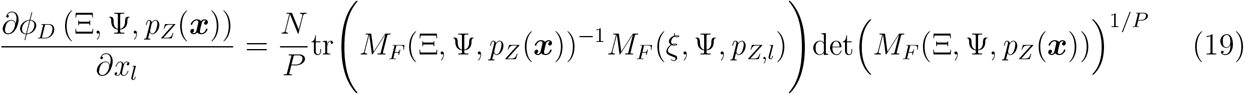

#### 2.4.4. Sample size computation

After having optimised the proportion of each covariate distribution, we can compute the expected standard errors on the parameters as the square roots of the diagonal elements of the inverse of the FIM. From that, we can derive the sample size needed, *NSN*, all other things being equal, to reach desired confidence level on parameter estimates or on power of statistical tests on covariate effect. Denoting *SE*(Ξ, Ψ, *p*_*Z*_(***x***))_*β*_ the standard error on a parameter *β* ∈ Ψ for a design Ξ, population vector Ψ and a covariate distribution defined by *p*_*Z*_ and ***x***, the number of subjects needed *NSN* to reach a desired level *SE*^⋆^ on this parameter is given in equation (20).

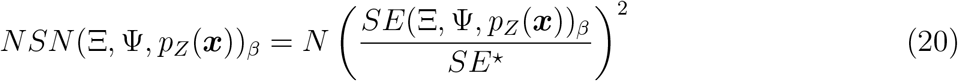

From this, we can calculate the *NSN* required to achieve the desired power in covariate testing.

### 2.5. Evaluation of covariate relationships

Usually, to assess covariate relationships, significance test are performed to determine whether covariate effects are different from 0. The expected power for this test can be computed from the FIM (Retout et al., 2007; Fayette et al., 2025).But to assess the magnitude of a covariate effect, one can compute the ratio of change in primary parameters when covariate values change relative to a reference value *z*_*ref*_. The uncertainty on this ratio can be derived from the SE on the covariate effect parameter. Then, the effect of the covariate value on the considered parameter will be said clinically relevant if the 90% confidence interval on the ratio is completely outside an equivalence interval [*R*_*inf*_; *R*_*sup*_] and clinically non-relevant if entirely within. If the confidence interval crosses the equivalence area, there is not enough power to conclude. Detailed formulas for ratios and powers are given in (Fayette et al., 2025). For a log-normally distributed parameter with an additive covariate relationship on the log scale, if the covariate is binary, the ratio writes 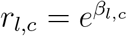. If continuous, the ratio associated to a given percentile *PX* of the covariate distribution writes 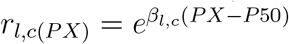, where *P* 50 denotes the median of the covariate distribution.

To assess the influence of belonging to an interval subset involving several covariates, we define the effect ratio as the expected value of the ratio function, conditional on the associated covariate sub-vector following the mixture distribution corresponding to the interval subset of interest.

Denoting by ℒ ^*R*^ the subset of covariate distributions defined over the interval subset, the ratio is given in Equation (21).

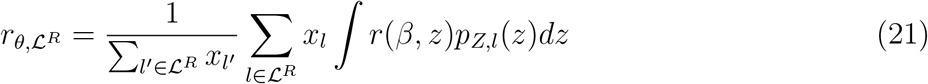

The uncertainty associated with this composite ratio can be estimated using the Delta Method, and both relevance and non-relevance tests can be conducted based on the resulting Gaussian approximation.

## 3. Illustration of benefit of covariate distribution optimisation on a one-compartment PK model

In this section we explored a PK example to demonstrate the benefit of covariate distribution optimisation in reducing the sample size needed to draw conclusions trough to case studies, the first one in the context of renal impairment and the second of hepatic impairment.

### 3.1. PK example

#### 3.1.1. Model

The PK model was a one compartment model with intravenous (IV) bolus and linear elimination, thus the drug concentration wrote 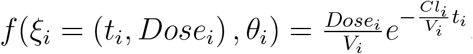, with two parameters: the clearance (*Cl*) and the volume of distribution (*V*), thus *θ*_*i*_ = (*Cl*_*i*_, *V*_*i*_)^⊤^. The residual error was modelled as a combined error model: *g*(*ξ*_*i*_, *θ*_*i*_, *σ* = (*a, b*)^⊤^) = *a* + *bf* (*ξ*_*i*_, *θ*_*i*_) and exponential random effects without correlation were assumed for *V* and 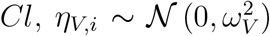 and 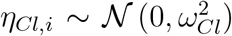. All covariate effects were modelled as additive on the log-scale. Both case studies include an effect of the Sex (*SEX*) and of the Body Mass Index (*BMI*) on *V*. *BMI* was first normalised and log transformed: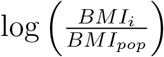, where *BMI*_*pop*_ denoted the reference *BMI* among reference individuals.

##### Renal case study

In the first case study an effect of RF on *Cl* was added through the creatinine clearance (CLCR), which was also normalised and log transformed: 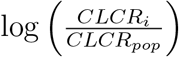, where *CLCR*_*pop*_ denoted the reference *CLCR* among reference individuals.

##### Hepatic case study

In the second case study effect of HF was added on *Cl*, based on serum total Bilirubin (*BILI*) and serum Aspartate transaminase (*AST*) concentrations (Ramalingam et al., 2010), which were also normalised and log transformed: 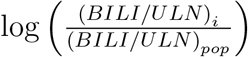 and 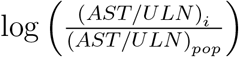, where (*BILI/ULN*)_*pop*_ and (*AST/ULN*)_*pop*_ denote the references among reference individuals and *ULN* stands for Upper Limit Normal (ULN).

#### 3.1.2. Covariate distribution specification

To reflect the case where covariates are not available at the design stage, we used the public database from the NHANES database (Centers for Disease Control and Prevention (CDC).National Center for Health Statistics (NCHS), 2009-2020) to get prior covariate distributions. Covariate data from 2009 to 2020 were extracted, keeping only subjects between 18 and 80 years old, with *BMI* higher or equal to 18.5 *kg/m*^2^ and for whom there were no missing data for Age, *SEX*, Body Weight (BW), *BMI*, Height, Creatinine (*CR*_*serum*_), *BILI* and *AST*. 29 409 covariate vectors were selected in the NHANES database and Table 1 provides the summary statistics of the covariates.

**Table 1:** Covariate data summary.

| Continuous covariates |  |  |
| --- | --- | --- |
| Covariate | mean (sd) | median [min; max] |
| *BMI* (kg/m<sup>2</sup>) | 29.5 (7) | 28.2 [18.5; 92.3] |
| *CLCR* (ml/min) | 99 (35.6) | 97.3 [4.1; 342.9] |
| *BILI/ULN* | 0.6 (0.3) | 0.6 [0; 7.3] |
| *AST/ULN* | 0.7 (0.5) | 0.7 [0.2; 26.7] |
| Discrete covariates |  |  |
| Covariate | Category Name | N (%) |
| *SEX* | Male | 14 340 (48.8%) |
|  | Female | 15 069 (51.2%) |
| Continuous segmentation |  |  |
| Condition on covariate | Interval Name | N (%) |
| $18.5 \leq BMI < 25 \text{ kg/m}^2$ | Healthy Weight | 8 217 (27.9%) |
| $25 \leq BMI < 30 \text{ kg/m}^2$ | Overweight | 9 620 (32.7%) |
| $BMI \geq 30 \text{ kg/m}^2$ | Obesity | 11 572 (39.3%) |
| $CLCR \geq 90 \text{ mL/min}$ | Normal RF | 17 187 (58.4%) |
| $60 \leq CLCR < 90 \text{ mL/min}$ | Mild RF | 8 327 (28.3%) |
| $30 \leq CLCR < 60 \text{ mL/min}$ | Moderate RF | 3 536 (12%) |
| $CLCR < 30 \text{ mL/min}$ | Severe RF | 359 (1.2%) |
| $BILI/ULN \leq 1$ and $AST/ULN \leq 1$ | Normal HF | 24 714 (84%) |
| $BILI/ULN \leq 1$ and $AST/ULN > 1$ | Mild1 HF | 2 597 (8.8%) |
| $1 < BILI/ULN \leq 1.5$ | Mild2 HF | 1 739 (5.9%) |
| $1.5 < BILI/ULN \leq 3$ | Moderate HF | 349 (1.2%) |
| $3 < BILI/ULN$ | Severe HF | 10 (0%) |

*BMI*: refers to Body Mass Index, *CLCR*: Creatinine Clearance, *RF*: Renal function, *BILI*: Bilirubin, *AST*: Aspartate transaminase, *ULN*: Upper Limit Normal, *HF*: Hepatic function

The body weight was adjusted for ideal body weight computed with Robinson’s formula (Robinson et al., 1983) and the Cockcroft and Gault equation (equation (22)) with *BW*_*Adj*_ the adjusted body weight (Chin et al., 2013) was then used to estimate the creatinine clearance, *CLCR*.

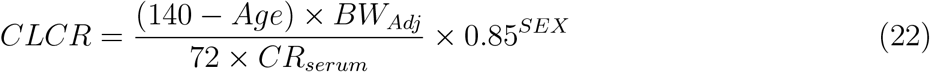

Both *BILI* and *AST* were normalised by their respective ULN, which were 1.0 mg/dL for *BILI* and 33 U/L for *AST* before 2017, and then 37 U/L for Male and 31 U/L for Female.

The *BMI* was segmented into three usual intervals, namely Obesity if *BMI* ≥ 30 *kg/m*^2^, Overweight if 25 ≤ *BMI <* 30 *kg/m*^2^ and Healthy Weight if 18.5 ≤ *BMI <* 25 *kg/m*^2^. Of note, *BMI*_*pop*_ = 22.8 *kg/m*^2^, *CLCR*_*pop*_ = 112.3 *mL/min*, (*BILI/ULN*)_*pop*_ = 0.6 and (*AST/ULN*)_*pop*_ = 0.64.

##### Renal case study

*CLCR* was segmented into the four usual intervals to define RF, namely Normal RF if *CLCR* ≥ 90 *mL/min*, Mild RF if 60 ≤ *CLCR <* 90 *mL/min*, Moderate RF if 30 ≤ *CLCR <* 60 *mL/min* and Severe RF if *CLCR <* 30 *mL/min*. While there was a good balance between Male and Female and between the different *BMI* intervals, Severe RF was poorly represented with only 1.2% of the data. Combining *SEX, BMI* intervals and *CLCR* intervals, there were 2 *×* 4 *×* 3 = 24 possible covariate combinations. Nevertheless, those including Severe RF were poorly represented in the dataset (e.g. only 40 subjects are Male, with Severe RF and Obesity). Therefore, Severe RF individuals were pooled into two classes depending on their *SEX* and without taking into account their *BMI* status. This pooling was carried out to ensure the adequacy of copula fits. The detailed repartition between the resulting 20 covariate distributions is given in the first panel of Figure 8.

##### Hepatic case study

HF was characterized using the National Cancer Institute (NCI) Organ Dysfunction Working Group (ODWG) criteria (Ramalingam et al., 2010). According to these criteria, Normal HF is defined as *BILI/ULN* ≤ 1 and *AST/ULN* ≤ 1. Mild HF includes two subgroups: defined as *BILI/ULN* ≤ 1 and *AST/ULN >* 1 or 1 *< BILI/ULN* ≤ 1.5 with any *AST/ULN*. For simplicity, we refer to these subpopulations as Mild1 and Mild2. Moderate and Severe HF are defined by 1.5 *< BILI/ULN* ≤ 3 and *BILI/ULN >* 3, respectively. Due to the very limited representation of Severe HF (only 10 subjects), Moderate and Severe HF were pooled, together accounting for only 1.2% of the covariate database. Consequently, as with RF, the ModerateSevere HF category was only combined with *SEX*. We chose to split the Mild HF category and model its distribution as a mixture of two copulas, each fitted independently to the Mild1 and Mild2 subgroups. As a result, a total of 20 copulas were fitted, while only 14 proportions required optimisation. The detailed distribution across the resulting 14 covariate distribution is shown in the first panel of Figure 11.

Reference individuals were defined as Male with Healthy Weight and both Normal RF and HF.

#### 3.1.3. Parameter values

The PK parameter values and ratios of covariate effects are given in Table 2. With a ratio of 0.70, i.e. outside of [0.80; 1.25], the effect of being a Female on *V* was tested for relevance; while *r*_*V*, Obesity_ was within the equivalence interval, thus the effect of Obesity on *V* was tested for non-relevance. It should be noted that *r*_*V*, Obesity_ was computed for the threshold value for belonging to this interval, namely for 30 *kg/m*^2^, in relation to the reference value, *BMI*_*pop*_.

**Table 2:** Population PK parameter values used in the illustration example.

| Parameter | $\mu_{Cl}$ (L/h) | $\mu_V$ (L) | $\omega_{Cl}$ | $\omega_V$ | $a$ (mg/L) | $b$ |
| --- | --- | --- | --- | --- | --- | --- |
| Value | 0.43 | 7.13 | 0.24 | 0.30 | 2.39 | 0.08 |

Covariate effects
| Parameter | $\beta_{V,Sex}$ | $\beta_{V,BMI}$ | $\beta_{Cl,CLCR}$ | $\beta_{Cl,BILI/ULN}$ | $\beta_{Cl,AST/ULN}$ |
| --- | --- | --- | --- | --- | --- |
| Value | -0.35 | 0.50 | 0.27 | 0.05 | -0.38 |

Ratios
| Ratio | $r_{V, Female}$ | $r_{V, Obesity}$ | $r_{Cl, Severe RF}$ | $r_{Cl, ModerateSevere HF}$ |
| --- | --- | --- | --- | --- |
| Value | 0.70 | 1.14 | 0.70 | 0.97 |

$\mu_*$ : refers to the typical value, $\beta_*$ : covariate parameter, $\omega_*$ : standard deviation of the random effect, $a$ and $b$ , parameters of the residual error model
$r_{l,c}$ : ratio on the parameter $l$ for the covariate $c$

##### Renal case study

The ratio assessing the impact of Severe RF on *Cl* was computed at the threshold value of 30 *ml/min*, relative to the reference *CLCR*_*pop*_. The resulting ratio was 0.70, which lies outside the equivalence interval [0.80; 1.25], and was therefore tested for relevance.

A sensitivity analysis was conducted on this example to evaluate two aspects. First, the influence of IIV was assessed using two scenarios: a ‘Normal IIV’ scenario, with standard deviations as specified in Table 2, and a ‘High IIV’ scenario, in which these standard deviations were doubled. Second, the impact of covariate values was explored by varying *β*_*Cl,CLCR*_, resulting in the ratio *r*_*Cl*, Severe RF_ ranging from 0.40 to 0.75.

##### Hepatic case study

Through this case study we wanted to explore a non-relevant relationship, therefore small effect sizes were chosen with *β*_*Cl,BILI/ULN*_ = 0.05 and *β*_*Cl,AST/ULN*_ = −0.38. The ratio assessing the impact of ModerateSevere HF depends on both *BILI/ULN* and *AST/ULN*, and was computed as the expected value over covariate distributions involving ModerateSevere HF, as described in Section 2.5. The resulting ratio was 0.97, which falls within the equivalence range.

#### 3.1.4. Design

The study design includes 100 subjects, each receiving a 250 mg dose of the drug at time 0. For each subject, three blood samples are collected at 1, 4, and 12 hours post-dose. The concentration-time profiles, *f* (*t*), accounting for univariate covariate effects and parameter values provided in Table 2, are shown in Figure 1. Because being Female reduces the volume of distribution, the typical concentration curve for Females lies above that for Males. Similarly, since a higher BMI increases the volume, the typical concentration curve for individuals with a Healthy Weight is higher than that for Overweight individuals, which in turn is higher than that for those with Obesity. Regarding RF, lower *CLCR* values correspond to lower concentration curves, reflecting reduced clearance due to renal impairment. HF is more complex, as it depends on two covariates. When considered independently, higher *BILI/ULN* values are associated with lower concentration curves, while higher *AST/ULN* values correspond to higher concentrations.

**Figure 1.**
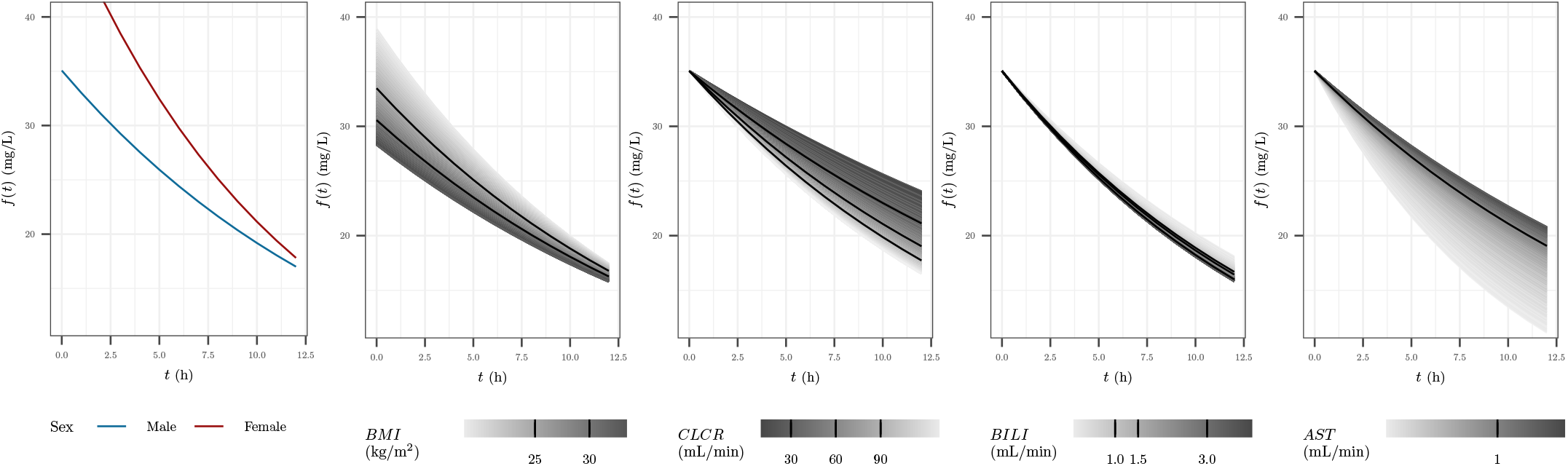
Evolution of the concentration *f* (*t*) according to the fixed effects and univariate covariate effects: the first panel corresponds to the effect of *SEX* on *V*, the second panel *BMI* on *V*, the third panel *CLCR* on *Cl*, the fourth panel *BILI/ULN* on *Cl* and the fifth panel *AST/ULN* on *Cl*. For for the last four panels, the black lines correspond to the limits of the intervals of the continuous covariates.

### 3.2. FIM evaluation using copula and quadrature

#### 3.2.1. Copula fitting

In both case studies, all copula were fitted as vine copula, on the associated subset of data, using the R package *rvinecopulib*0.6.3.1.1 (Nagler and Vatter, 2021). Copula fit evaluation was conducted through a simulation-based strategy as described in (Guo et al., 2024b) using 100 replicates. The relative error (RE) between summary metrics *M*_*sim*_ in copula-simulated populations and *M*_*obs*_ in the NHANES dataset was computed as 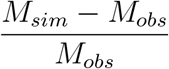. Summary metrics were the mean, the standard deviation, the median and the 5^*th*^ and 95^*th*^ percentiles of the marginal distribution of each continuous covariate. Overlaps between simulated and true distributions were also computed such as the simulated and observed correlations between continuous covariates. Diagnostics plots were explored using the R package *pmxcopula* (Guo et al., 2024a) *with minor edits*.

#### 3.2.2. Quadrature settings

According to the GLQ rule, the approximation using *n*^(*Q*)^ nodes is exact for integrands that are polynomials of degree 2*n*^(*Q*)^ − 1 or less. Nevertheless, in our case, the integrand is the composition of the FIM and the inverse Rosenblatt transform of the copula describing the continuous covariate distribution, therefore, in a general setting, it is not a polynomial function. Consequently, the quadrature remains an approximation and increasing *n*^(*Q*)^ increases the accuracy. To determine the number of nodes required in the quadrature for an acceptable accuracy in FIM evaluation, we computed the FIM starting with *n*^(*Q*)^ = 1 and going up to *n*^(*Q*)^ = 15, and looked for stabilization in the D-criterion by computing the relative difference in 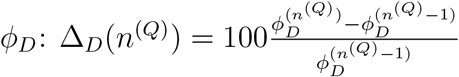 between two number of nodes. We stopped when the latter was bellow 0.5%. The same number of nodes was used for all the continuous covariates and for each copula. As a target, we also computed the FIM using Monte Carlo simulations with *n*^(*MC*)^ = 500 Monte Carlo samples drawn from each copula and because of the stochasticity of Monte Carlo, we repeated this process 10 times and defined the target D-criterion as the average across the 10 repetitions.

### 3.3. Covariate distribution optimisation

In all scenarios, covariate distribution optimisation was carried using PGD. The step-size *α* was was fixed at 0.001. Optimisation was terminated when the change in the objective function between two successive iterations was less than *ϵ* = 10^−5^. Two constraint settings were considered. The first, referred to as ‘Without constraints’, imposed only the basic requirements that the proportions lie within the interval [0, 1] and sum to 1. The second setting, referred to as ‘With constraints’, introduced additional constraints: each continuous interval was required to represent at least 5% of the population. Furthermore, the Severe RF and ModerateSevere HF were capped to 10% in each case study respectively. Indeed, such subjects are difficult to recruit in real-world practice.

### 3.4. Optimised distributions evaluation

The initial distribution was the observed proportions in the selected subset of the NHANES database. For each optimised distribution, the D-efficiency was calculated relative to the design based on the initial distribution. Additionally, the *NSN* to achieve 80% power for detecting the relevance or non-relevance of the relationships was computed.

### 3.5. Implementation

All the methods have been implementing using the R language (R Core Team, 2021). For FIM evaluation an extension of the package *PFIM* 6.1 (Mentré et al., 2024), based on previous work (Fayette et al., 2025) have been done. All scripts are available online at https://doi.org/10.5281/zenodo.14778033. The quadrature nodes were computed using the function *gauss*.*quad()* from the package *statmod* (Giner and Smyth, 2016). For PGD, the projection on the constraint space was made using the function *lsei()* from the package *limSolve* (Karline Soetaert et al., 2009). *Roots for NSN* were found using the R function *uniroot()* from the package *stats*(R Core Team, 2013) which implements algorithms from (Brent, 2013).

## 4. Results

### 4.1. Copula fitting

The performance of copula model was evaluated for each covariate combination.

#### 4.1.1. Renal case study

As shown in Appendix A, Figure A.13, RE was within ±10% for all the summary statistics for almost all the intervals of *BMI* and *CLCR*. The simulated correlations from the copula models were very similar to observed correlations between *BMI* and *CLCR* (Appendix A, Figure A.14) and overlap between simulated and observed join distribution for *BMI* and *CLCR* was always above 80%, except for Female with Severe RF, for which the overlap was above 80% in only in three quarters of the simulations (Appendix A, Figure A.15). Moreover, the VPC given in Figure 2 showed a good match between simulated and observed level lines, for all combinations. Overall, copula fits were judged to be very satisfactory.

**Figure 2.**
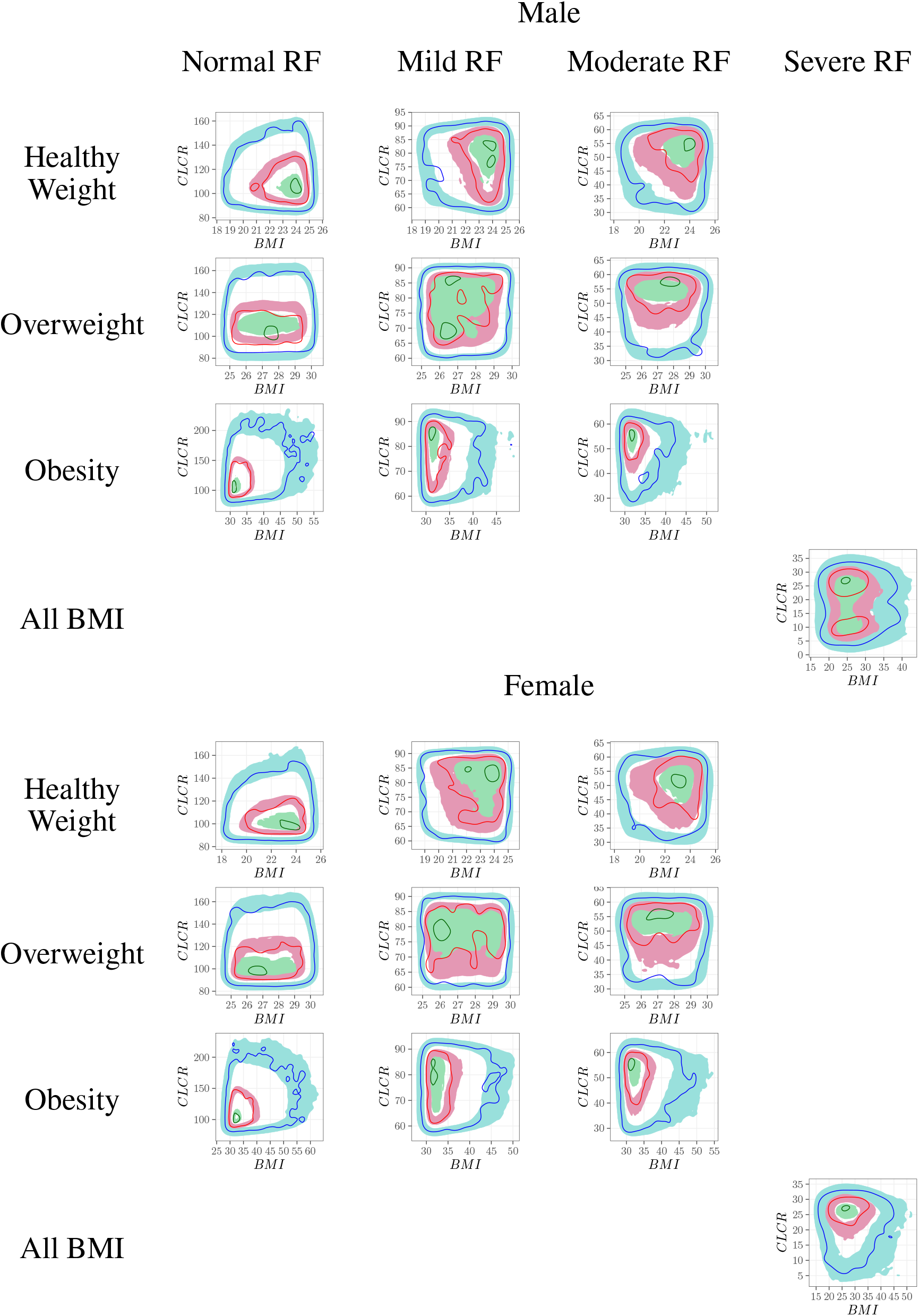
Renal case study - Visual predictive checks based on the contours of the bivariate density between *BMI* and *CLCR*. For each of the 20 covariate combination, the virtual population was simulated 100 times from the copula and the 99% prediction intervals of percentile contours of the joint distribution was derived and compared to the contours observed in the NHANES database. The ribbon areas correspond to the 99% prediction interval of 5^*th*^ 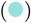, 50^*th*^ 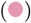and 95^*th*^ 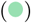 percentile contours of the joint distribution of *BMI* and *CLCR*. The lines corresponds to the observed 5^*th*^ 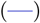, 50^*th*^ 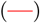 and 95^*th*^ 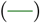 percentile contours.

#### 4.1.2. Hepatic case study

Regarding the hepatic case study, the overall fit quality appeared to be somewhat lower, particularly in terms of marginal estimations for *AST/ULN*. Specifically, the relative error (Appendix A, Figure A.16) seemed to be less well controlled. Nevertheless, the correlation structure between variables was adequately captured (Appendix A, Figure A.14). The overlap (Appendix A, Figure A.15) was not very satisfactory for the subgroup of females with ModerateSevere HF, which can be attributed to the limited number of observed subjects in the dataset (only 77). Importantly, however, the VPC (Figure 3) did not reveal any anomalies.

**Figure 3.**
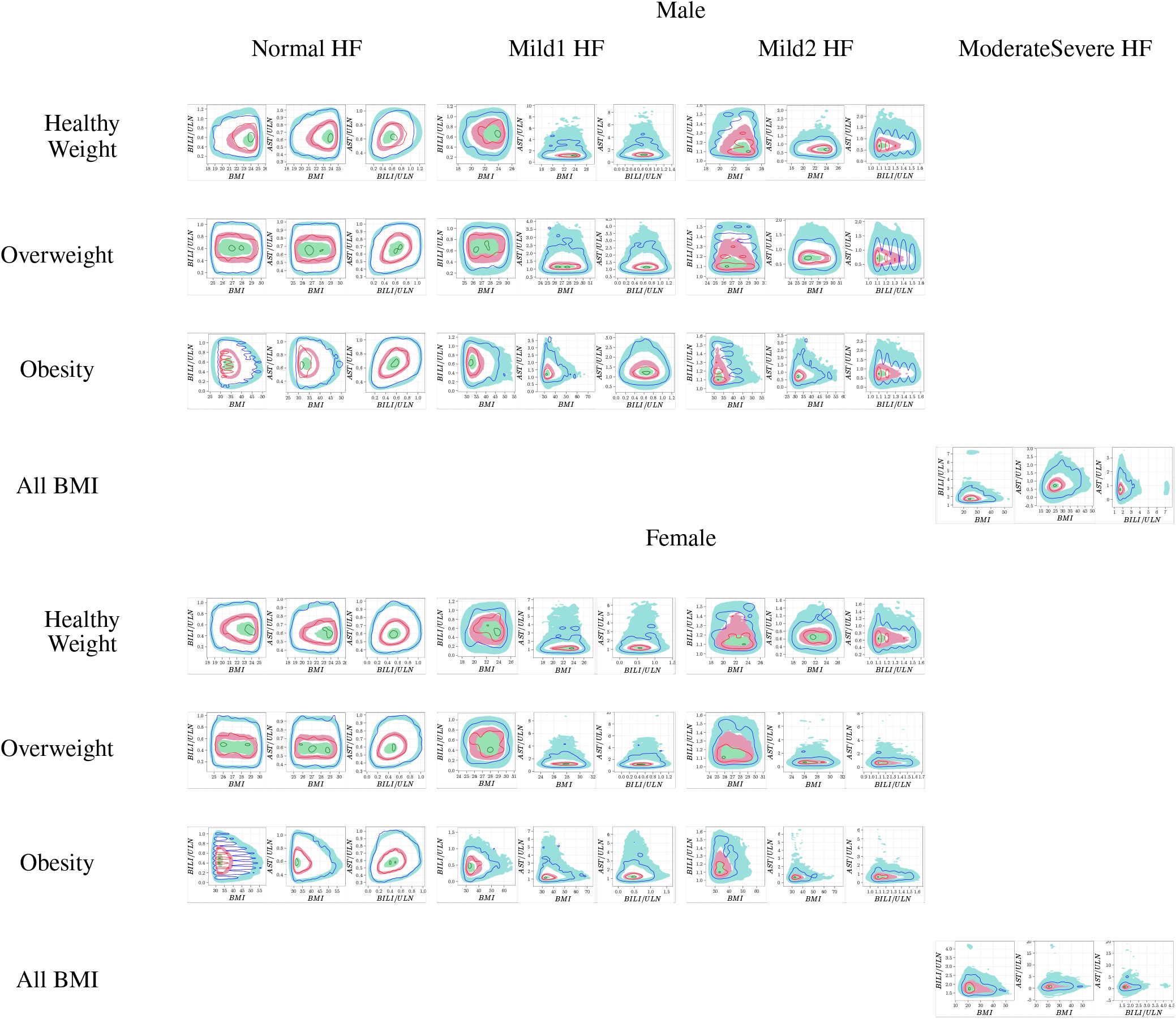
Hepatic case study - Visual predictive checks based on the contours of the bivariate density between *BMI, BILI/ULN* and *AST/ULN*. For each of the 20 covariate combination, the virtual population was simulated 100 times from the copula and the 99% prediction intervals of percentile contours of the joint distribution was derived and compared to the contours observed in the NHANES database. The ribbon areas correspond to the 99% prediction interval of 5^*th*^ 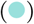, 50^*th*^ 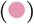 and 95^*th*^ 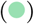 percentile contours of the joint distributions. The lines corresponds to the observed 5^*th*^ 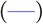, 50^*th*^ 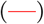 and 95^*th*^ 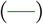 percentile contours.

### 4.2. FIM evaluation using quadrature

#### 4.2.1. Renal case study

Figure 4 presents the convergence in relative difference of the D-criterion. The relative difference in *ϕ*_*D*_ between two successive number of nodes dropped below 0.5% at *n*^(*Q*)^ = 5, yielding a Dcriterion value of 258.60. Using Monte Carlo integration (Figure 5) with *n*^(*MC*)^ = 500 repeated 10 times, the target D-criterion was 260.1 (sd = 0.8), corresponding to a difference of 0.6% with the GLQ of 5 nodes. However, GLQ required 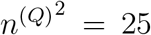 FIM evaluations for each covariate distributions, while for Monte Carlo, the relative error compared to the target was not controlled *±* 0.5% for *n*^(*MC*)^ = 25 (Figure 5).

**Figure 4.**
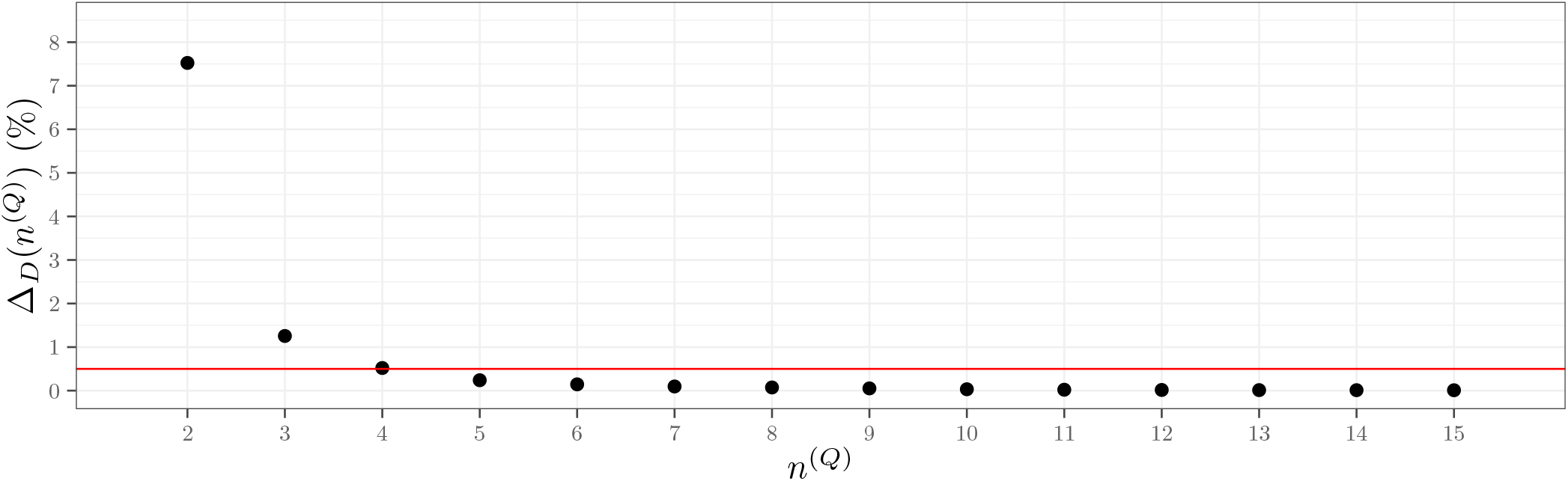
Renal case study - Relative difference in D-criterion as a function of the number of nodes, *n*^(*Q*)^, in the Gauss-Legendre Quadrature used for Fisher Information Matrix integration

**Figure 5.**
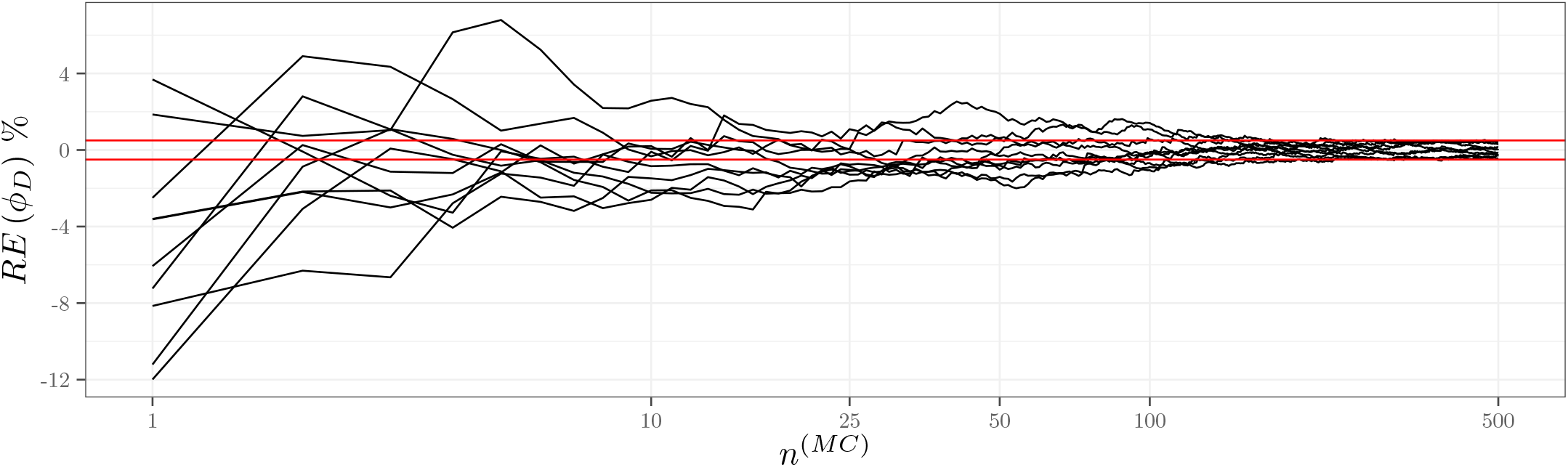
Renal case study - Relative error compared to the target D-criterion computed as the average across 10 repetitions of FIM integration using *n*^(*MC*)^ = 500 Monte Carlo samples as a function of the number of Monte Carlo samples used in Fisher Information Matrix integration The red line corresponds to the target *±*5%.

Moreover, as shown in Appendix B Figure B.19, the convergence behaviour for GLQ was consistent across the two IIV scenarios, regardless of the ratio value. Therefore, the value *n*^(*Q*)^ = 5 was selected for the optimisation step.

#### 4.2.2. Hepatic case study

In the HF case study, the relative difference in the overall D-criterion also dropped below 0.5% at *n*^(*Q*)^ = 6, yielding a D-criterion value of 240.1 (Figure 6). Using Monte Carlo integration (Figure 7) with *n*^(*MC*)^ = 500 repeated 10 times, the target D-criterion was 243.8 (sd = 0.9), corresponding to a difference of 1.5%, deemed acceptable and *n*^(*Q*)^ = 6 was kept for optimisation. Of, note, the convergence was not monotonic in this case study, but remained controlled under a difference of 0.5% from *n*^(*Q*)^ = 6.

**Figure 6.**
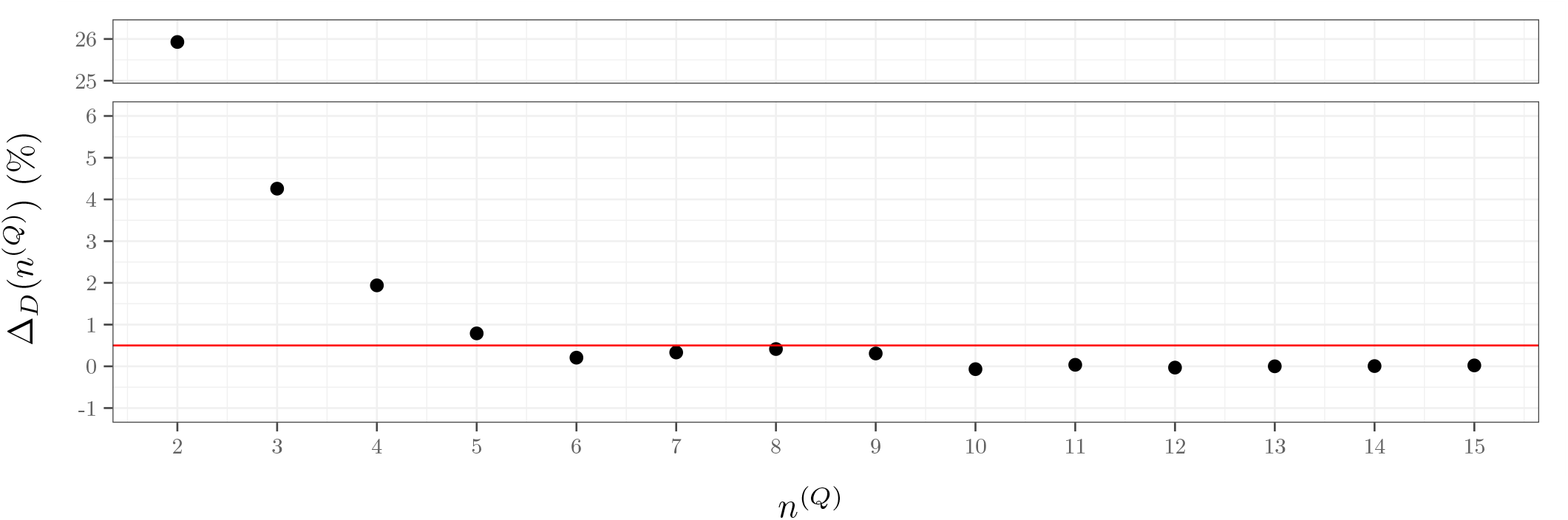
Hepatic case study - Relative difference in D-criterion as a function of the number of nodes, *n*^(*Q*)^, in the Gauss-Legendre Quadrature used for Fisher Information Matrix integration The red line corresponds to the 0.5%.

**Figure 7.**
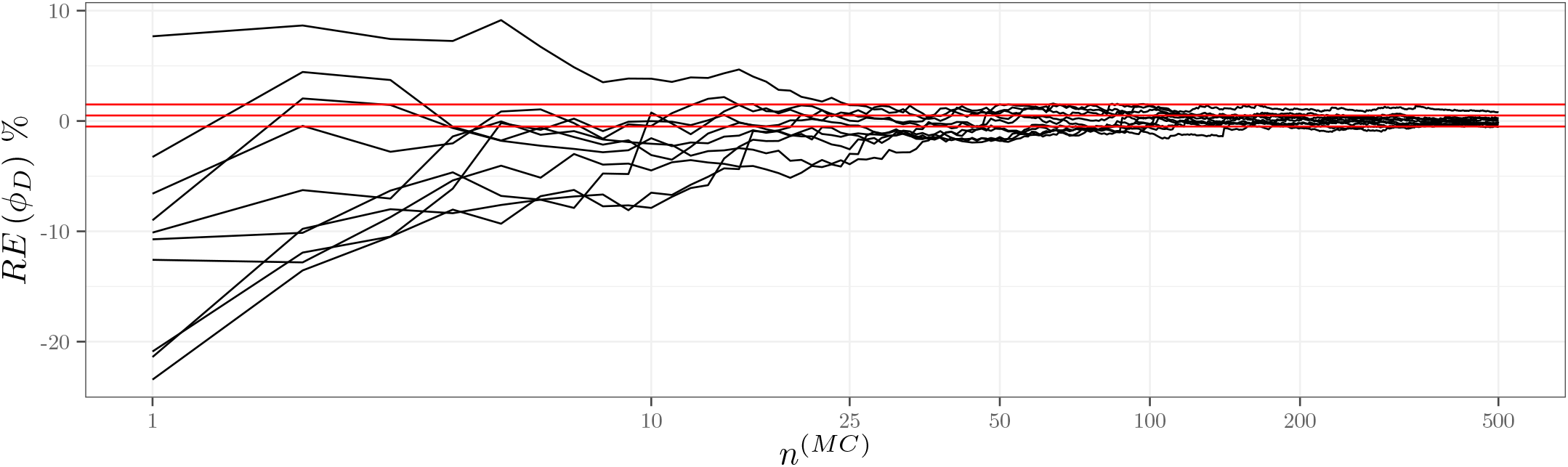
Hepatic case study - Relative error compared to the target D-criterion computed as the average across 10 repetitions of FIM integration using *n*^(*MC*)^ = 500 Monte Carlo samples as a function of the number of Monte Carlo samples used in Fisher Information Matrix integration The red line corresponds to the target *±*5%.

### 4.3. Covariate distribution optimisation

In both case studies, the PGD converged without trouble and convergence plots are given in Appendix C, Figures C.20 and C.22.

#### 4.3.1. Renal case study

The distribution between the 20 covariate combinations in the RF case study are given in Figure 8 for the initial distribution and for the two optimisation settings. The first observation is that the optimal distributions were very sparse in the sense that many categories had a weight of 0.

**Figure 8.**
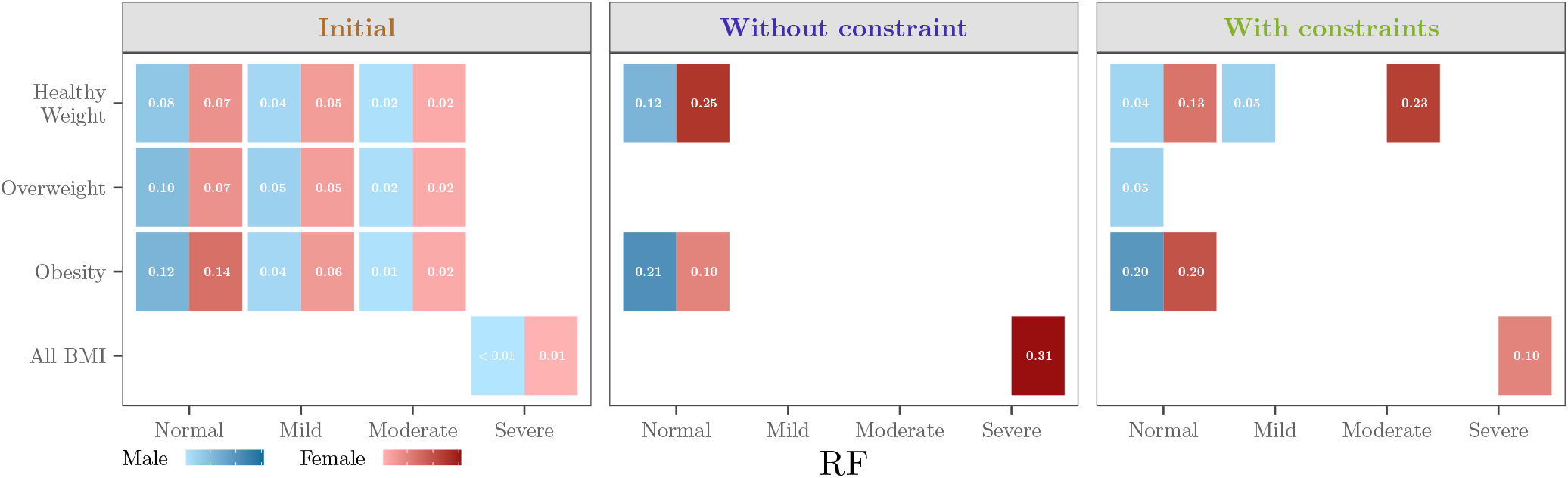
Renal case study - Optimal proportions for the covariate distributions for the D-criterion for the different sets of constraints

Because being a Female reduces *V* while *V* increases with the *BMI*, in optimal distributions, Female were more associated with Healthy Weight while Male were associated with Obesity. Similarly, because having a Severe RF decreases *Cl* and therefore slows the elimination and leads to higher concentrations, in optimal distributions, only Female Severe RF were present.

Overall, the general shape of the optimal distributions remained relatively stable across different levels of IIV and *CLCR* effect (Appendix C Figure C.21). However, it can be observed that with increasing IIV or decreasing covariate effect magnitude, greater weight was assigned to more renal impaired.

Marginal distributions are provided in Appendix C, Table C.5. For *SEX*, the optimal allocation consistently favoured females, with 66% representation, regardless of the constraint settings. For *BMI* intervals, Overweight individuals appeared in the optimal distribution, even without explicit constraints, due to the pooling of *BMI* categories within the Severe RF combinations. The proportions of Healthy Weight and Obesity were nearly unchanged between the unconstrained and constrained scenarios, 47% and 43% without constraints, versus 49% and 43% with constraints, respectively. Regarding RF, without constraint, Mild and Moderate RF were absent from the optimal distribution, resulting in a Normal–Severe RF split of 69% and 31%. With constraints, Mild RF was included at its minimum allowable level, and due to the additional upper constraint, Severe RF were at 10% and 23% were reallocated to Moderate RF.

Regarding performance, both optimisation settings improved D-efficiency (Table 3), by 23% without constraints and by 19% with constraints. Even under this more realistic constrained setting, the *NSN* was substantially reduced. Specifically, the highest *NSN* required to conclude on the relevance of being Female on *V*, of having Severe RF on *Cl*, and the non-relevance of Obesity on *V* was 602 with the initial design. This number dropped to respectively 197 and 230 without and with constraints. Thus, even the more constrained optimisation reduced the *NSN* by over 60%. The corresponding forest plot is shown in Figure 9 where it is evident that the initial distribution would not have allowed for a conclusion on relevance.

**Table 3:**
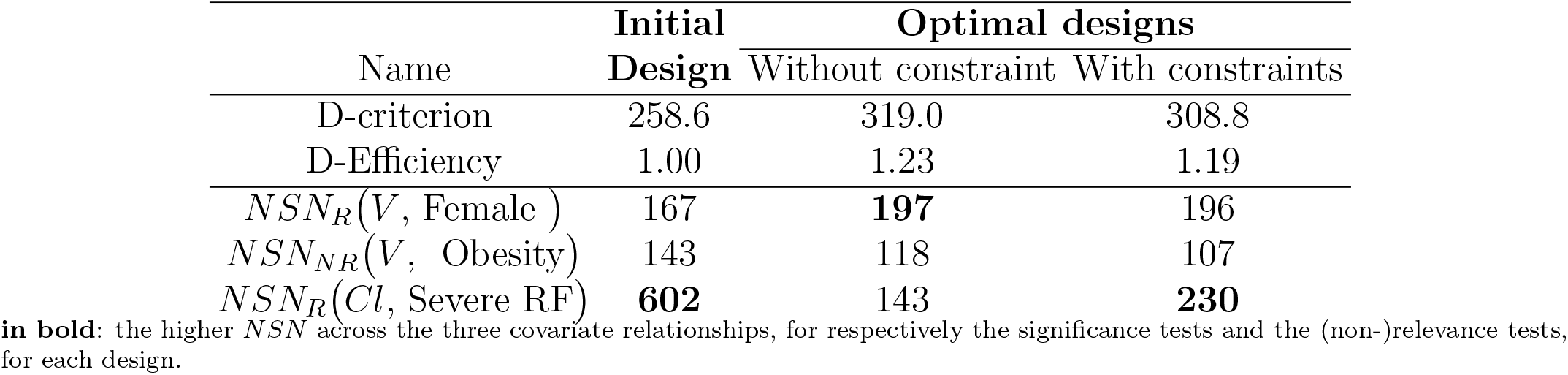
Renal case study - D-criterion, D-Efficiency and Number of subjects needed (*NSN*) to reach 80% power in relevance or non-relevance tests, with the initial and optimal covariate distributions for the D-criterion for the different sets of constraints

| Name | Initial | Optimal designs |  |
| --- | --- | --- | --- |
|  | Design | Without constraint | With constraints |
| D-criterion | 258.6 | 319.0 | 308.8 |
| D-Efficiency | 1.00 | 1.23 | 1.19 |
| $NSN_R(V, \text{Female})$ | 167 | **197** | 196 |
| $NSN_{NR}(V, \text{Obesity})$ | 143 | 118 | 107 |
| $NSN_R(Cl, \text{Severe RF})$ | **602** | 143 | **230** |

**in bold:** the higher $NSN$ across the three covariate relationships, for respectively the significance tests and the (non-)relevance tests, for each design.

**Figure 9.**
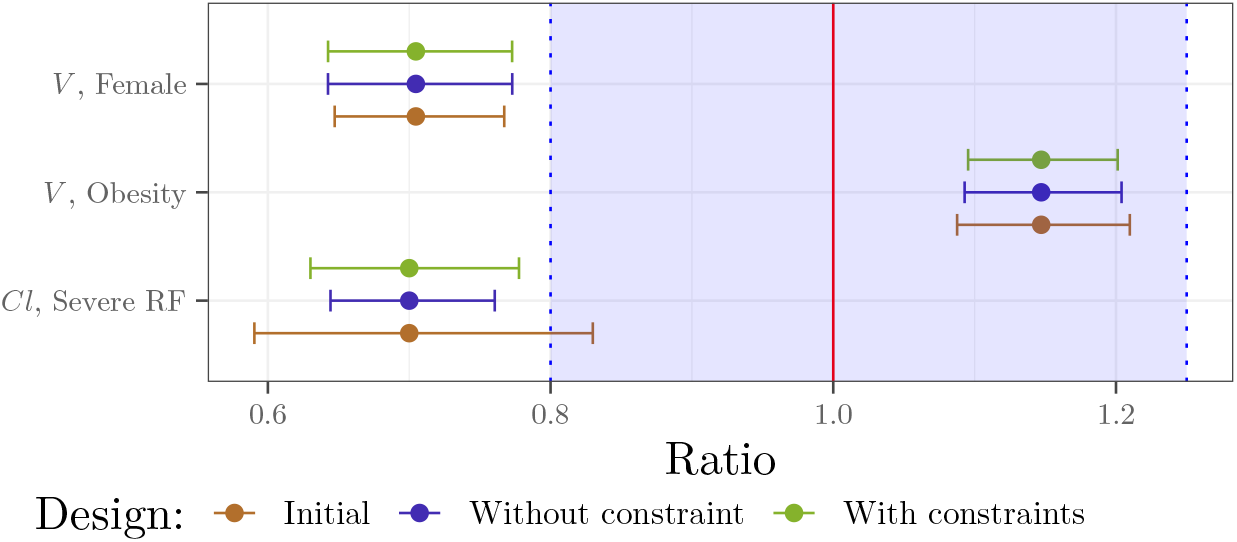
Renal case study - Ratios of covariate effects and their 90% confidence interval predicted with the Fisher Information Matrix for *N* = 230 subjects, using the initial covariate distribution and the optimal ones, without and with constraints. The red line at 1 corresponds to the reference line i.e., no change from the typical individual; the shaded area in blue represents the reference area of [0.80, 1.25].

Regarding sensitivity analysis, as expected the greater the effect of RF (i.e. the smaller the ratio), the smaller the *NSN* to conclude on relevance of Severe RF effect (Figure 10). For a same effect magnitude, the *NSN* increased with IIV.

**Figure 10.**
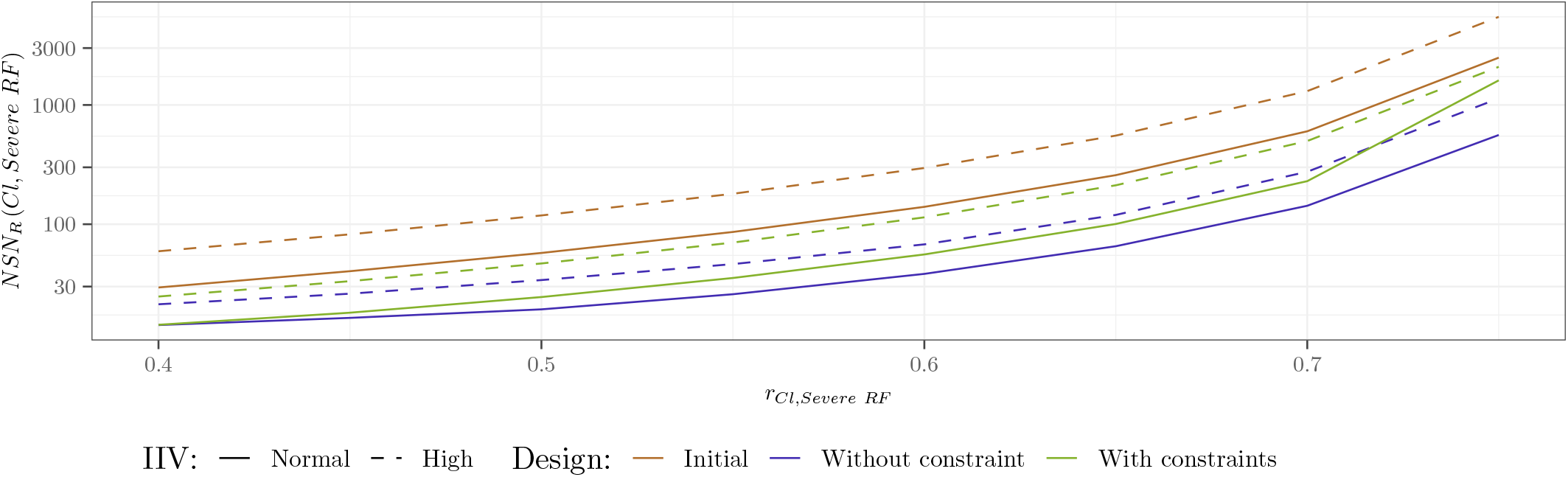
Renal case study - *NSN*_*R*_ *Cl, CLCR* (Severe RF) to reach 80% power in relevance test (right) for the two IIV scenarios and as a function of *r*_*Cl*, Severe RF_ from 0.40 to 0.75

#### 4.3.2. Hepatic case study

For the HF case study, the optimal distributions are shown in Figure 11. As in RF case study, the optimal allocation primarily weighted extreme values, with no Overweight individuals included in the unconstrained setting. As above, because ModerateSevere HF decreases *Cl*, Females were more associated with Healthy Weight and ModerateSevere HF, while Males were associated with Obesity and Normal HF. Notably, the optimal distributions included a higher proportion (72%) of Females than Males (Appendix C, Table C.6). Similarly to the RF case study, the unconstrained optimal marginal distribution for HF was mainly concentrated between Normal HF (44%) and ModerateSevere HF (40%). In the constrained setting, where a 10% upper bound was imposed on ModerateSevere HF, part of this group was replaced by Mild HF which became the majority.

**Figure 11.**
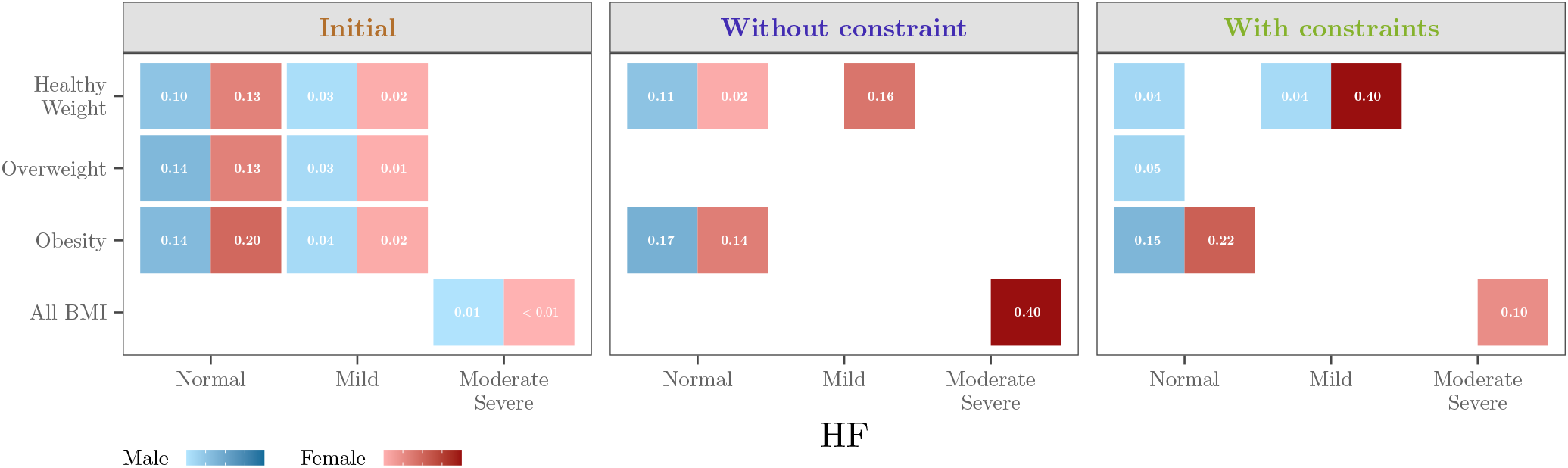
Hepatic case study - Optimal proportions for the covariate distributions for the D-criterion for the different sets of constraints

Regarding performances, optimisation once again increased the D-criterion and reduced the *NSN* (Table 4). The *NSN* to conclude on the relevance of the effect of being a Female on *V* and of the effect of having ModerateSevere HF on *Cl*, and on the non-relevance of having Obesity on *V* was 400 with the initial design. This number dropped to 216 with the constrained optimisation, representing a reduction of 46%. The corresponding forest plot is shown in Figure 12.

**Table 4:** Hepatic case study - D-criterion, D-Efficiency and Number of subjects needed (*NSN*) to reach 80% power in relevance or non-relevance tests, with the initial and optimal covariate distributions for the D-criterion for the different sets of constraints

| Name | Initial Design | Optimal designs |  |
| --- | --- | --- | --- |
|  |  | Without constraint | With constraints |
| D-criterion | 240.1 | 304.0 | 294.4 |
| D-Efficiency | 1.00 | 1.27 | 1.23 |
| $NSN_R(V, SEX)$ | 167 | **217** | **216** |
| $NSN_{NR}(V, Obesity)$ | 140 | 105 | 108 |
| $NSN_{NR}(Cl, ModerateSevere HF)$ | **400** | 148 | 141 |

in bold: the higher $NSN$ across the three covariate relationships, for respectively the significance tests and the (non-)relevance tests, for each design.

**Figure 12.**
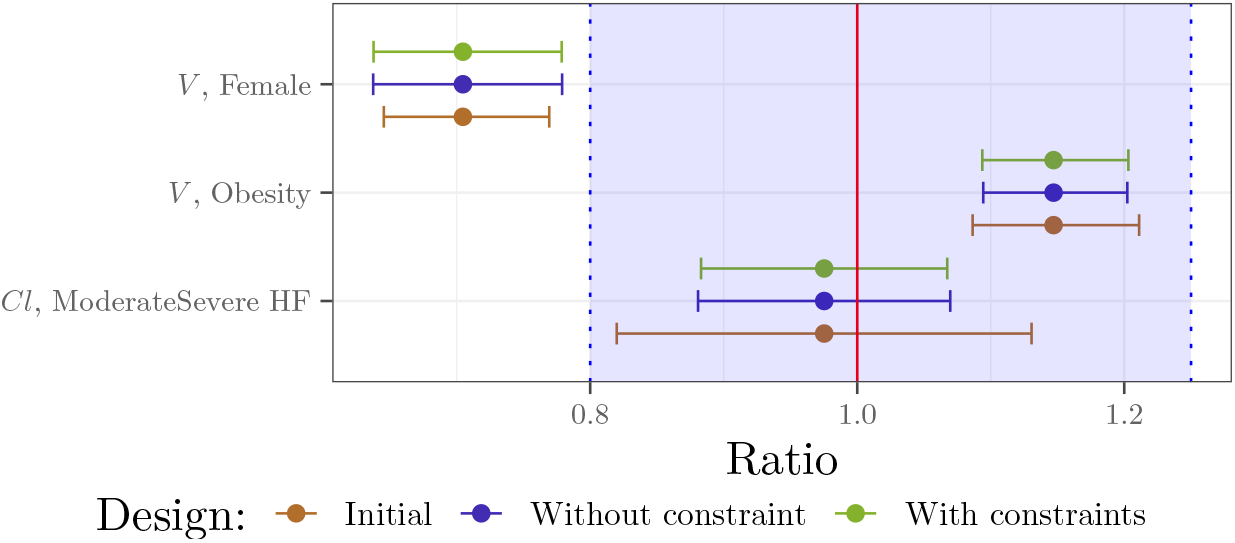
Hepatic case study - Ratios of covariate effects and their 90% confidence interval predicted with the Fisher Information Matrix for *N* = 216 subjects, using the initial covariate distribution and the optimal ones, without and with constraints. The red line at 1 corresponds to the reference line i.e., no change from the typical individual; the shaded area in blue represents the reference area of [0.80, 1.25].

## 5. Discussion

In this work we proposed to optimise the allocation of the covariates among the patients to be including in a future population PK study to assess the relevance or non-relevance of covariate relationships using NLMEM. We applied the proposed workflow to a PK model through two case studies, assessing the effects of either renal or hepatic impairment. We have first introduced a new way of integrating the FIM for the covariate distribution by leveraging copula modelling and GLQ. The optimisation was carried out between categories for discrete covariates, while for continuous ones, we introduced a segmentation of their domains into intervals and optimised the proportion between them. We solved the optimisation problem using the PGD algorithm. All these methods have been implemented in R based on the PFIM 6.1 package, and the source code is available online at the following link: https://doi.org/10.5281/zenodo.14778033.

In our example we used a covariate database on which copula were fitted as initial distributions. Goodness of fit diagnosis suggested that a large amount of data is required for estimating copula but the latter could have been used directly if available, which is one of their great interests in pharmaceutical applications as they can be made public without threatening anonymity. In addition, the optimisation approach can be coupled with another method for FIM computation if preferred. Of note, in the HF case study, we chose to fit separately the Mild1 and and the Mild2 data, although diagonal copula have been studied in the literature (see, for instance, (de Amo et al., 2016; Bukovšek et al., 2024)), but this topic was beyond the scope of our work.

We have shown that using GLQ for FIM integration with respect to covariate distribution provided good accuracy compared to Monte Carlo, while reducing the number of FIM evaluations and thus, computation time. We proposed a practical approach for users to determine the required number of quadrature nodes by monitoring the relative change in the D-criterion. If greater precision is required, the number of quadrature nodes can be increased; however, doing so may undermine the computational efficiency that motivates the use of quadrature methods compared to Monte Carlo. Indeed, the GLQ approach is limited by dimensionality: as the number of continuous covariates *C* increases, the number of FIM evaluations grows exponentially as 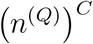. This challenge could be mitigated by refining the number of nodes per covariate, rather than using a uniform value or by employing adaptive quadrature methods that require fewer nodes (see, for example, the review in (Malcolm and Simpson, 1975)). That said, in practice it is unlikely that a large number of covariates would be optimized simultaneously as the objective is typically to evaluate only a few key covariates, as illustrated in our example. Moreover, GLQ remains computationally more efficient than CTS, which requires simulating and estimating a large number of datasets. If quadrature becomes computationally prohibitive, quasi–Monte Carlo methods (Pan and Thompson, 2007) may offer a practical compromise between standard Monte Carlo integration and deterministic quadrature, balancing accuracy and efficiency.

The introduced covariate segmentation enables a fast optimisation without the need to recompute FIM. This allows for the exploration of various constraints and optimisation settings without concern for computational cost. This optimisation was performed using the PGD with a absolute tolerance fixed at *ϵ* = 10^−5^ and a fixed step size. We did not explore these PGD hyper-parameters nor some of performance-improving variants because the need was not felt in our situation, but it can be an option in more complex cases. Especially, adaptive step size (Duchi et al., 2011) may have improved the solution or the computing time, which could be useful with higher dimensionality. In the current version of this algorithm, only linear constraints can be used for an easy projection on the constraint space, while in practice some non-linear ones may be useful (e.g., to impose the power of a test or a SE level on a parameter). To address this, a penalty term could be added to the objective function or another class of algorithms could be explored such as interior-point method (Lu and Pong, 2013). Regarding the optimisation criteria, we used the D-criterion and others exist such as the DS-or DDS-criterion which allow focus on a specific subset of the parameters (see for instance (Seurat et al., 2020) for an application). In our case, the covariates effects could have been the only parameters of interest. This change of criterion is not straightforward in the sense that its derivative has to be calculated, but apart from this calculation, it presents no particular difficulty with the PGD algorithm. The resulting optimal distributions may not correspond to integer subject counts within subgroups; however, efficient rounding methods (Pukelsheim and Rieder, 1992) can be applied to convert them into realistic sample sizes.

Regarding FIM computation, we used a linearisation approach but methods avoiding this approximation have already been explored using Monte Carlo and adaptive GQ (Riviere et al., 2016) (Ueckert and Mentré, 2017). A future work could explore a full evaluation of the FIM with no linearisation and covariate integration using these approaches. It would also allow extension to non-continuous response, for which the FIM cannot be computed with linearisation.

Last but not least, this approach assumes that the model and its parameters are known, including the effects of the covariates, whereas the trial to be designed aims to better estimate them, and is not robust to misspecifications. In the RF case study, we computed optimal allocation and *NSN* for a range of *β* and this could be extended into a robust approach as already explored in the NLMEM context (Dodds et al., 2005; Foo et al., 2012; Loingeville et al., 2020; Seurat et al., 2020). Moreover, this methodology could be used to design a Phase 3 study based on Phase 2 results; therefore, the SE from the covariate effects estimates based on the Phase 2 data may be used to specify a prior distribution on the *β*. This absence of robustness also holds for the covariate distribution assumed for FIM computation and covariate optimisation. In this case, Adaptive Designs and especially Two-stage approaches (Dodds et al., 2005; Lestini et al., 2015; Fayette et al., 2023) could be a good avenue to explore for refining the distribution of covariates halfway through inclusion.

From an application point of view, we illustrated our method through a PK example with two case studies in which we optimised the allocation of the covariates among the patients to be including in clinical trials to assess the relevance and non-relevance of renal and hepatic impairment. We especially focused on *NSN* to conclude on the relevance or non-relevance of the associated covariates with a sufficient degree of confidence. For this assessment, we used the equivalence region [0.80; 1.25] which is the one used in bioequivalence studies to define whether a dosing adjustment is required but alternative methods have been proposed to choose equivalence regions appropriate to a specific context (Sanghavi et al., 2024). In this work, we have assumed the same elementary design for all the subjects; the computations can easily be extended to the case where there are several. It would therefore certainly be more beneficial to adapt the elementary design to the covariate combination. Then, an interesting path may be to jointly optimize the elementary designs and the covariates. If discrete optimisation is performed for elementary designs, it can be combined with the covariates and the global process remains the same, only the number of FIM evaluations increases. We can imagine that this joint optimisation would enable us to improve the designs, for example in our case, for subjects with poor elimination, having a greater final observation time would guarantee a better estimation. On the other hand, for continuous optimisation, the extension is harder. In both cases, it will first be necessary to consider the practical and ethical constraints and discuss the actual implementation with experts. The resulting optimal distributions were consistent, typically favouring allocation to the more extreme classes, with intermediate values disappearing in scenarios without constraints. Constraints were introduced to better reflect real-world conditions for patient inclusion in clinical studies. Even with these constraints, our two case studies demonstrated that the optimal distribution greatly reduced the required *NSN* to conclude on the clinical relevance (or non-relevance) of the covariate relationships. The main challenge in implementing this optimal distribution lies in the need to recruit a substantial number of Female subjects with either Severe RF or ModerateSevere HF. Nevertheless, given the significant reduction in *NSN*, focusing efforts on these difficult-to-recruit subgroups may be justified. Moreover, constraints can be added to correspond practical settings. For instance we could have specified that we want as many Males with Severe RF (or ModerateSevere HF) as Females, or constraint each covariate combination to represent at least 5%. In addition, optimal proportions can be adapted to match actual conditions, but in this case, the *NSN* must be reevaluated using the new proportions.

Finally we introduced a flexible optimisation workflow to help the subject inclusion in clinical trials which aims to assess the relevance of covariates.

## Data Availability

All the R scripts are available at the following link: https://doi.org/10.5281/zenodo.16743214

## Code availability

All the R codes needed to reproduce the example in full are available online at: https://doi.org/10.5281/zenodo.14778033.

## Appendix A. Complementary results on copula diagnostics

### Appendix A.1. Renal case study

**Figure A13:**
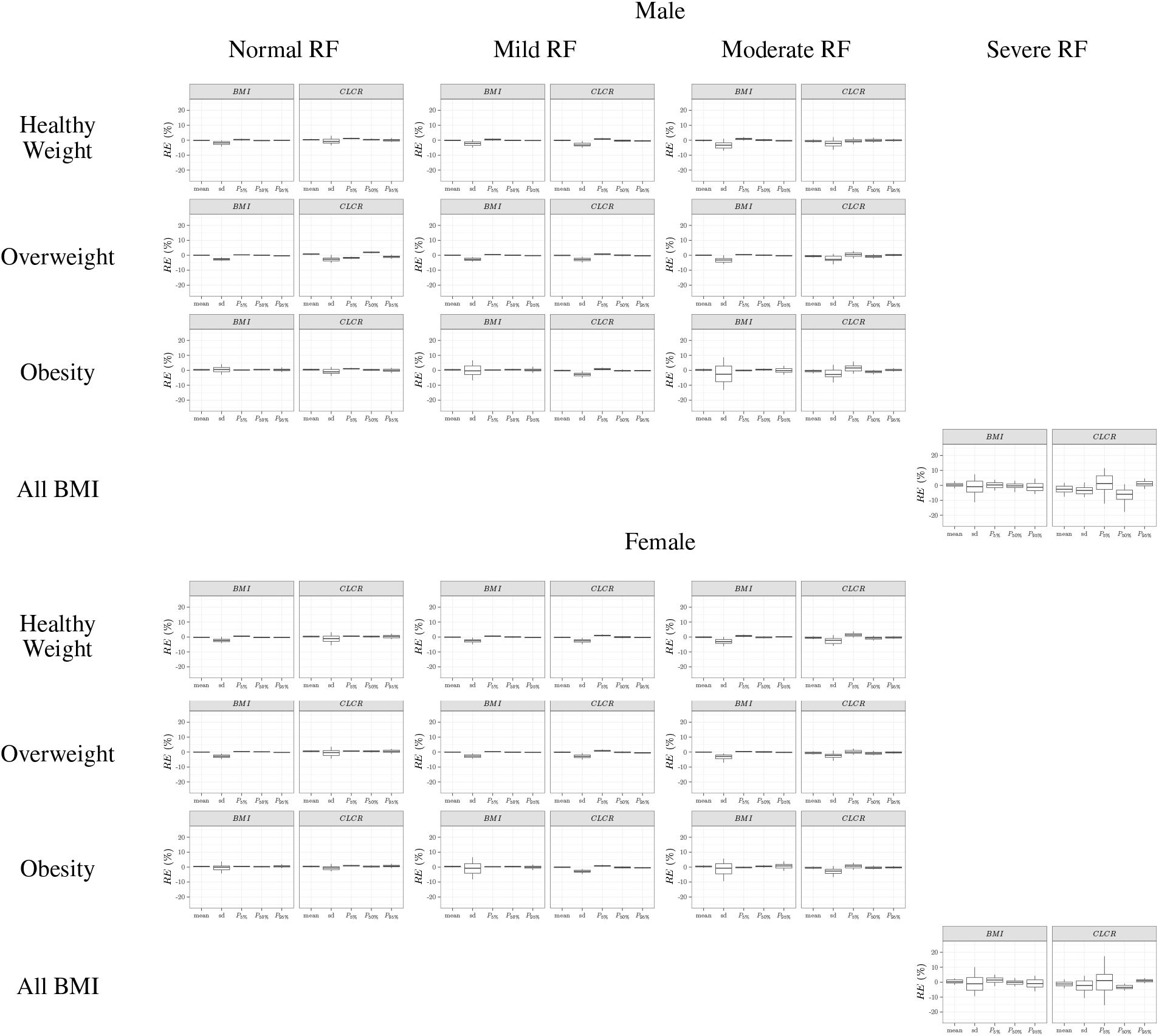
Renal case study - Boxplot of the relative error (*RE*) of marginal distributions (mean, standard deviation and percentiles) of continuous covariates for the 20 covariate combinations, as compared to the statistics of the NHANES database. For each combination, the virtual population was simulated 100 times from the copula. The boxplot displays the median, the 25^*th*^ and 75^*th*^ percentiles, while the whiskers are 5^*th*^ and 95^*th*^ percentiles. sd refers to the standard deviation, and *P* 5%, *P* 50% and *P* 95% to the 5^*th*^, 50^*th*^ and 95^*th*^ percentiles respectively.

**Figure A14:**
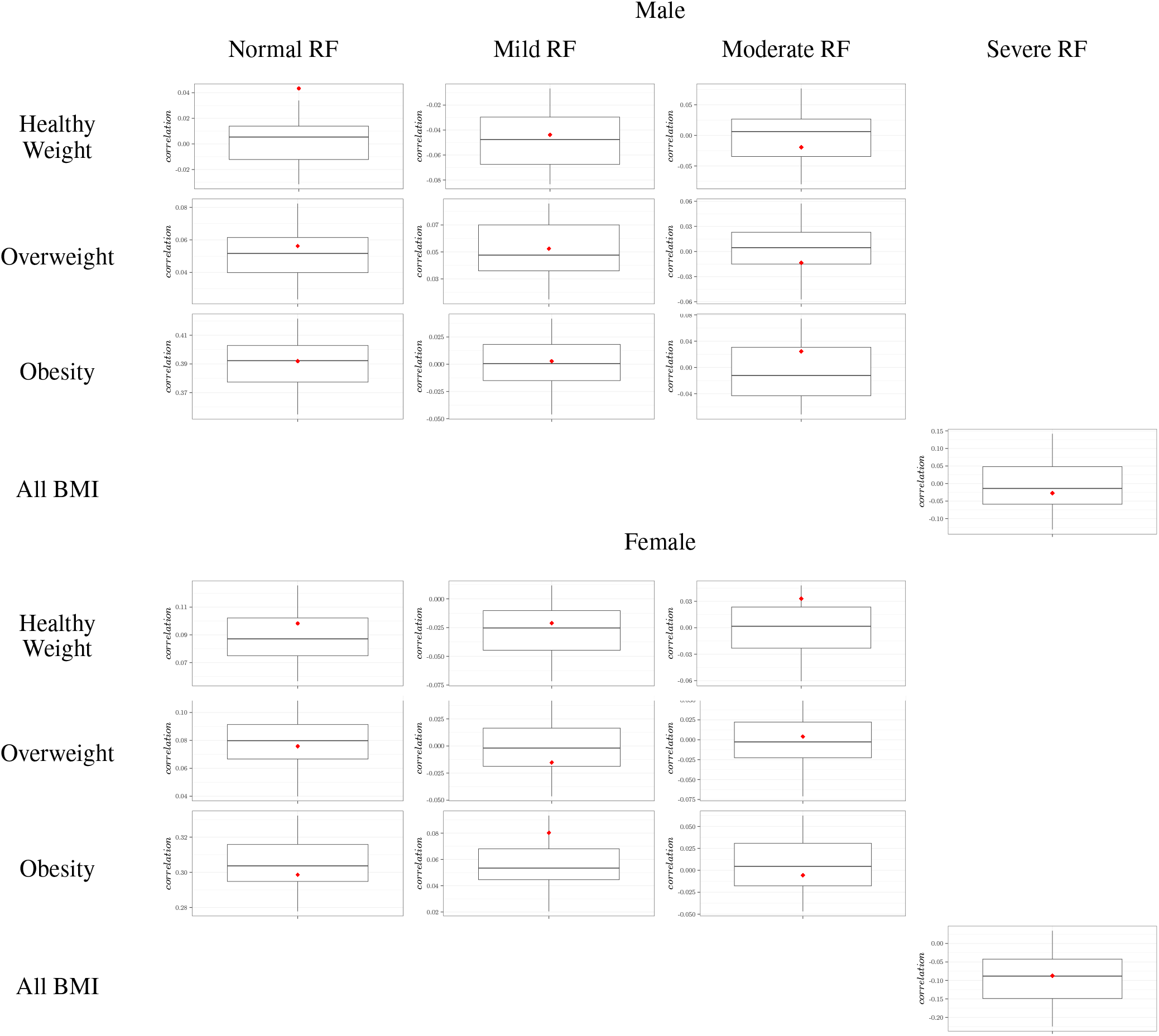
Renal case study - Correlation between *BMI* and *CLCR* for each of the 20 covariate combinations, in virtual population simulated 100 times from the copula (boxplot) and in the NHANES database (red diamond). The boxplot displays the median, the 25^*th*^ and 75^*th*^ percentiles, while the whiskers are 5^*th*^ and 95^*th*^ percentiles. sd refers to the standard deviation, and *P* 5%, *P* 50% and *P* 95% to the 5^*th*^, 50^*th*^ and 95^*th*^ percentiles respectively.

**Figure A15:**
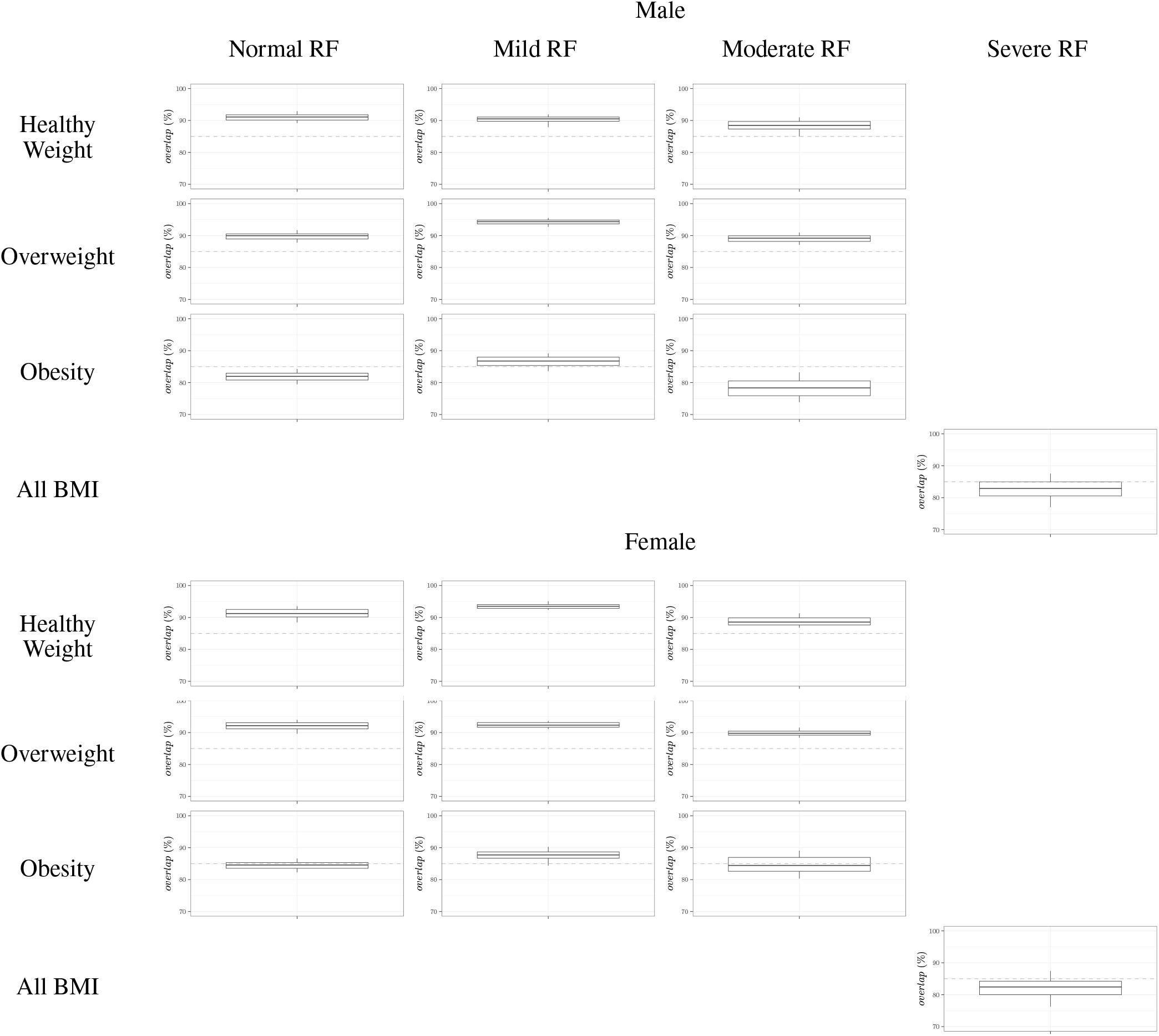
Renal case study - Overlap metric between the 95^*th*^ density contours of the joint distribution between *BMI* and *CLCR* in the virtual population simulated 100 times from the copula and in the NHANES database. Overlap metric of 95^*th*^ density contours of virtual population relative to observed population. The gray dashed line indicates 85% overlap percentage.

### Appendix A.2. Hepatic case study

**Figure A16:**
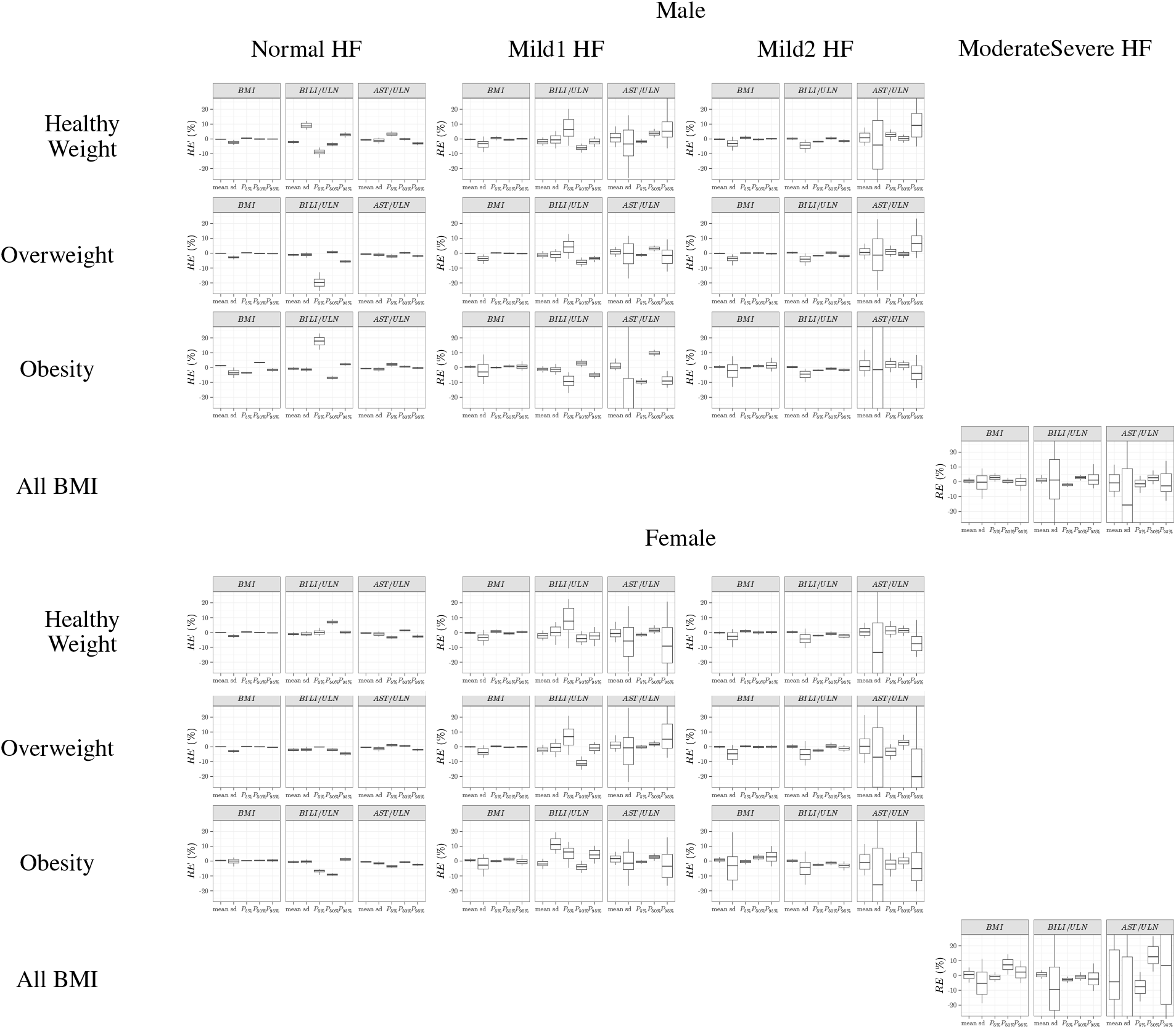
Hepatic case study - Boxplot of the relative error (*RE*) of marginal distributions (mean, standard deviation and percentiles) of continuous covariates for the 18 covariate combinations, as compared to the statistics of the NHANES database. For each combination, the virtual population was simulated 100 times from the copula. The boxplot displays the median, the 25^*th*^ and 75^*th*^ percentiles, while the whiskers are 5^*th*^ and 95^*th*^ percentiles. sd refers to the standard deviation, and *P* 5%, *P* 50% and *P* 95% to the 5^*th*^, 50^*th*^ and 95^*th*^ percentiles respectively.

**Figure A17:**
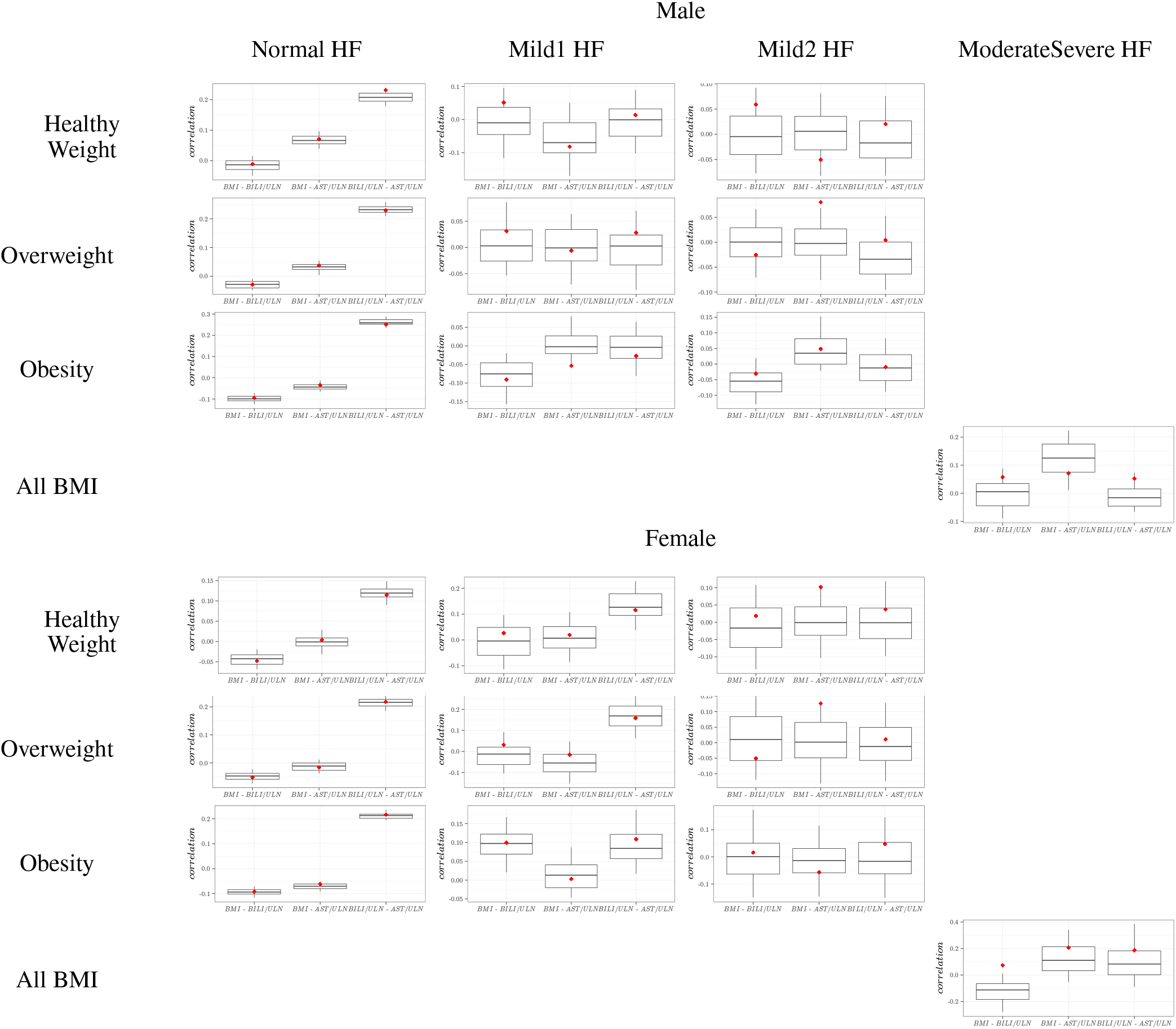
Hepatic case study - Correlations between *BMI, BILI/ULN* and *AST/ULN* for each of the 18 covariate combinations, in virtual population simulated 100 times from the copula (boxplot) and in the NHANES database (red diamond). The boxplot displays the median, the 25^*th*^ and 75^*th*^ percentiles, while the whiskers are 5^*th*^ and 95^*th*^ percentiles. sd refers to the standard deviation, and *P* 5%, *P* 50% and *P* 95% to the 5^*th*^, 50^*th*^ and 95^*th*^ percentiles respectively.

**Figure A18:**
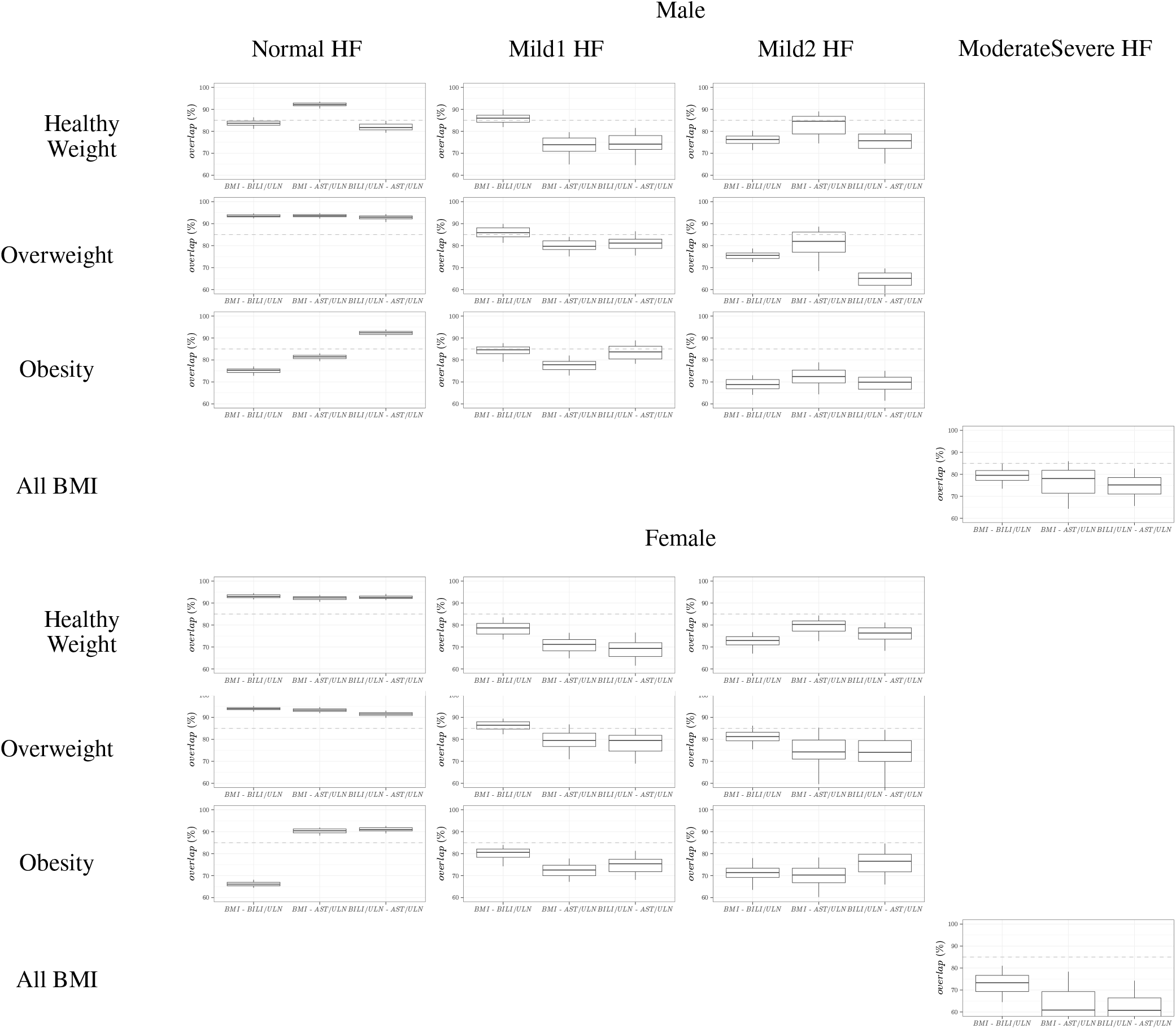
Hepatic case study - Overlap metric between the 95^*th*^ density contours of the joint distributions bbetween *BMI, BILI/ULN* and *AST/ULN* in the virtual population simulated 100 times from the copula and in the NHANES database. Overlap metric of 95^*th*^ density contours of virtual population relative to observed population. The gray dashed line indicates 85% overlap percentage.

## Appendix B. Complementary results on FIM evaluation using quadrature

**Figure B19:**
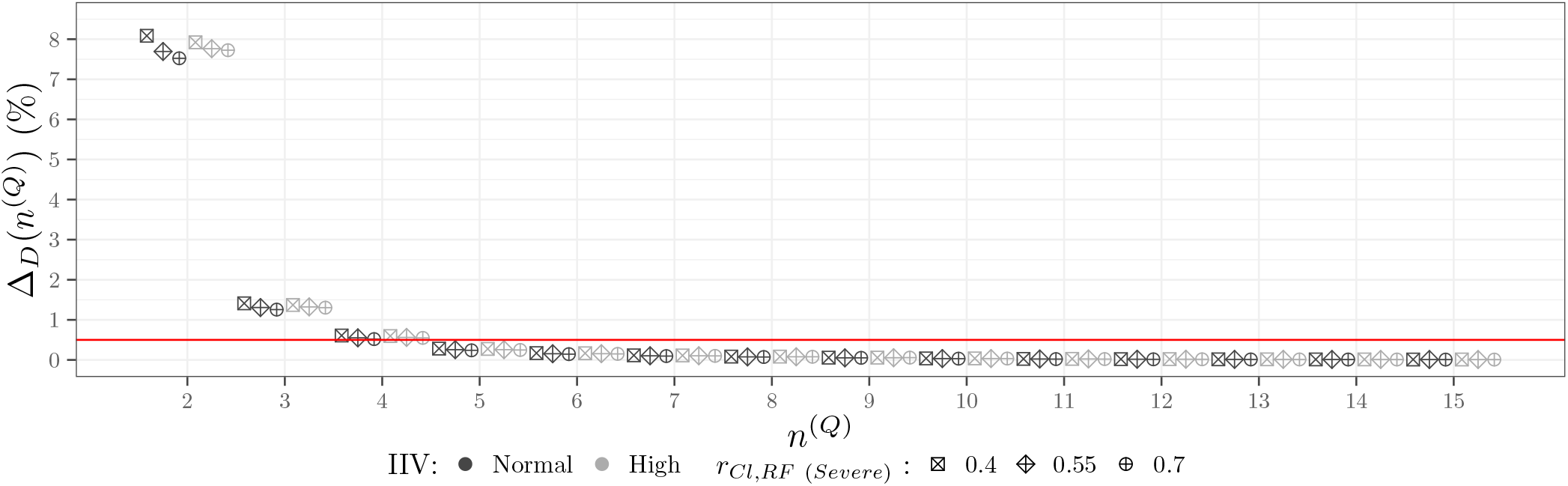
Renal case study - Sensitivity analysis for the relative difference in D-criterion as a function of the number of nodes, *n*^(*Q*)^, in the Gauss-Legendre Quadrature used for Fisher Information Matrix integration, for the two IIV scenarios and three values of *r*_*Cl*, Severe RF_ The red lines corresponds to the 0.5% target.

## Appendix C. Complementary results on covariate distribution optimisation

### Appendix C.1. Renal case study

**Figure C20:**
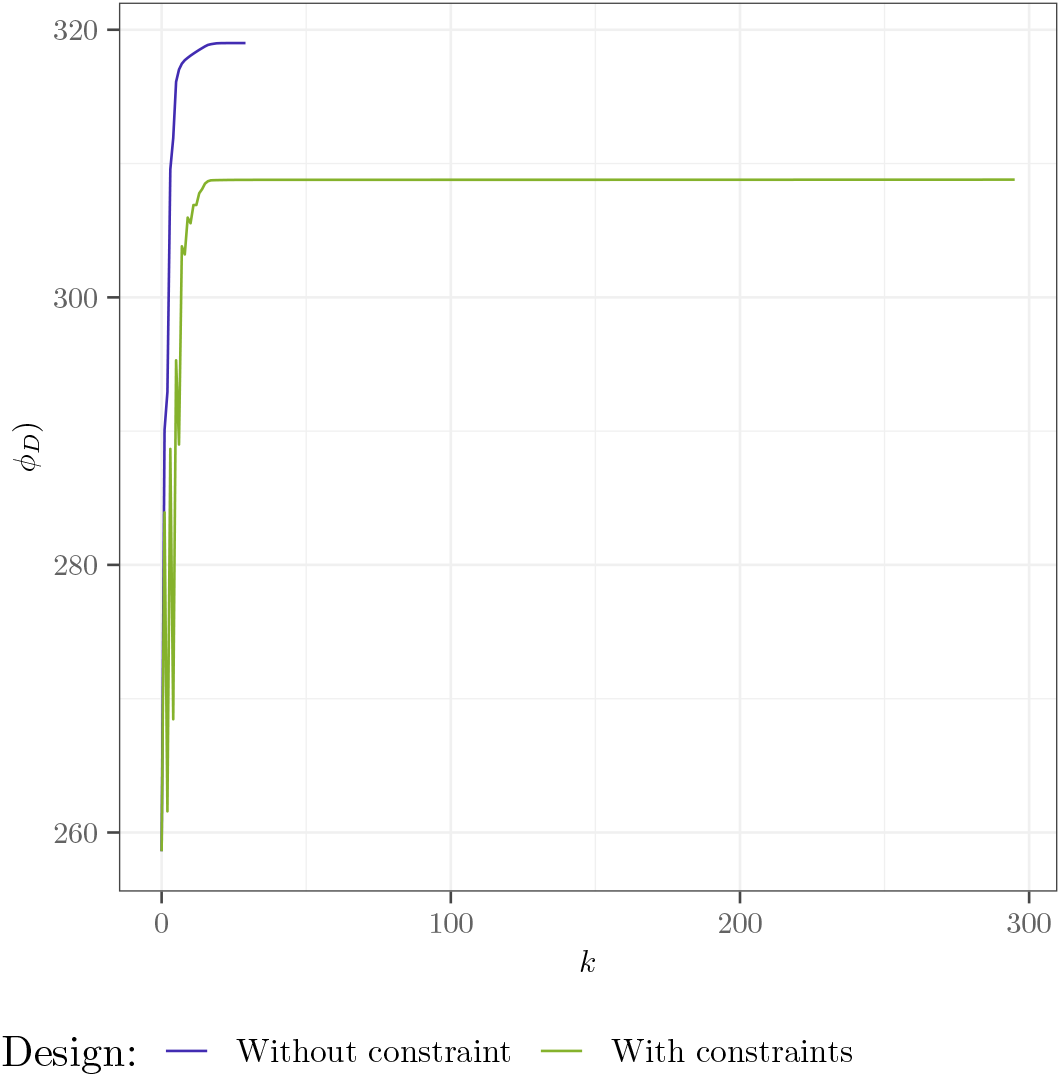
Renal case study - D-criterion as a function of the iterations in the Projected Gradient Descent optimisation for the different sets of constraints

**Figure C21:**
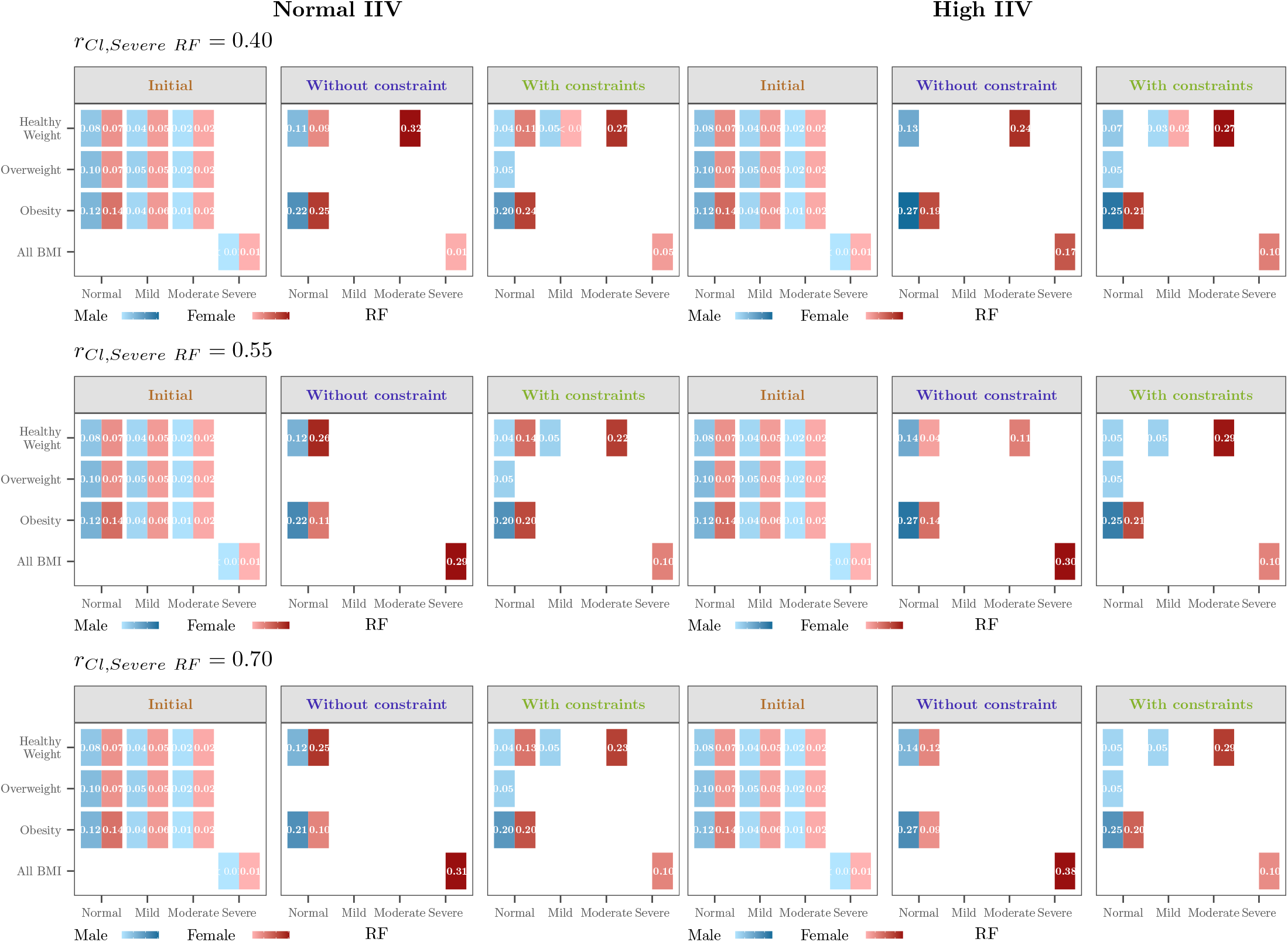
Renal case study - Optimal proportions for the covariate distributions for the D-criterion for the different sets of constraints, for the scenario with Normal IIV (left) and High IIV (right) and for three value of *r*_*Cl*, Severe RF_.

**Table C5:** Renal case study - Marginal proportions for the covariate distributions for the D-criterion for the different sets of constraints.

| Covariate | Interval | Initial | Without constraint | With constraints |
| --- | --- | --- | --- | --- |
| *SEX* | Male | 0.49 | 0.34 | 0.34 |
|  | Female | 0.51 | 0.66 | 0.66 |
| *BMI* | Healthy Weight | 0.28 | 0.47 | 0.49 |
|  | Overweight | 0.33 | 0.10 | 0.08 |
|  | Obesity | 0.39 | 0.43 | 0.43 |
| RF | Normal | 0.58 | 0.69 | 0.62 |
|  | Mild | 0.28 | 0.00 | 0.05 |
|  | Moderate | 0.12 | 0.00 | 0.23 |
|  | Severe | 0.01 | 0.31 | 0.10 |

### Appendix C.2. Hepatic case study

**Figure C22:**
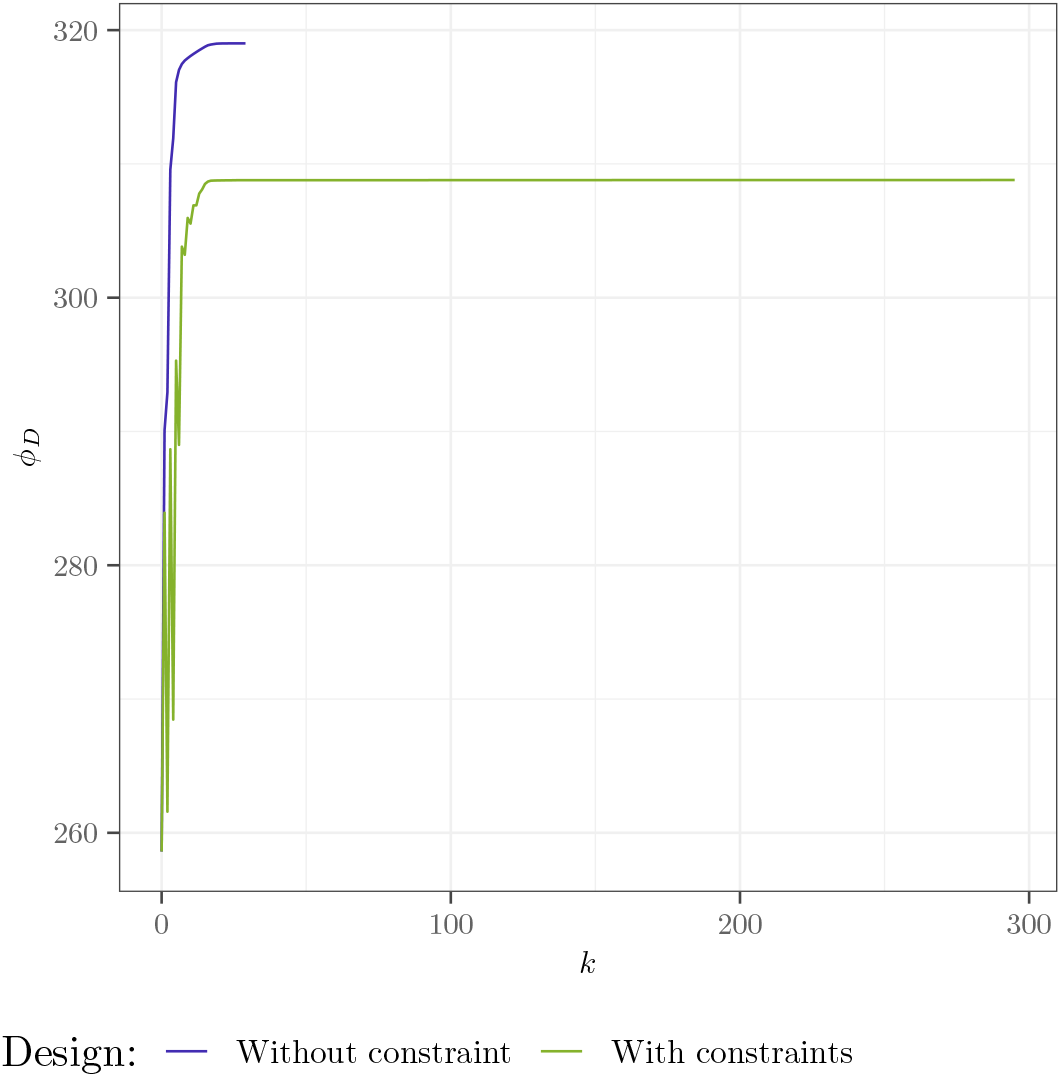
Hepatic case study - D-criterion as a function of the iterations in the Projected Gradient Descent optimisation for the different sets of constraints

**Table C6:**
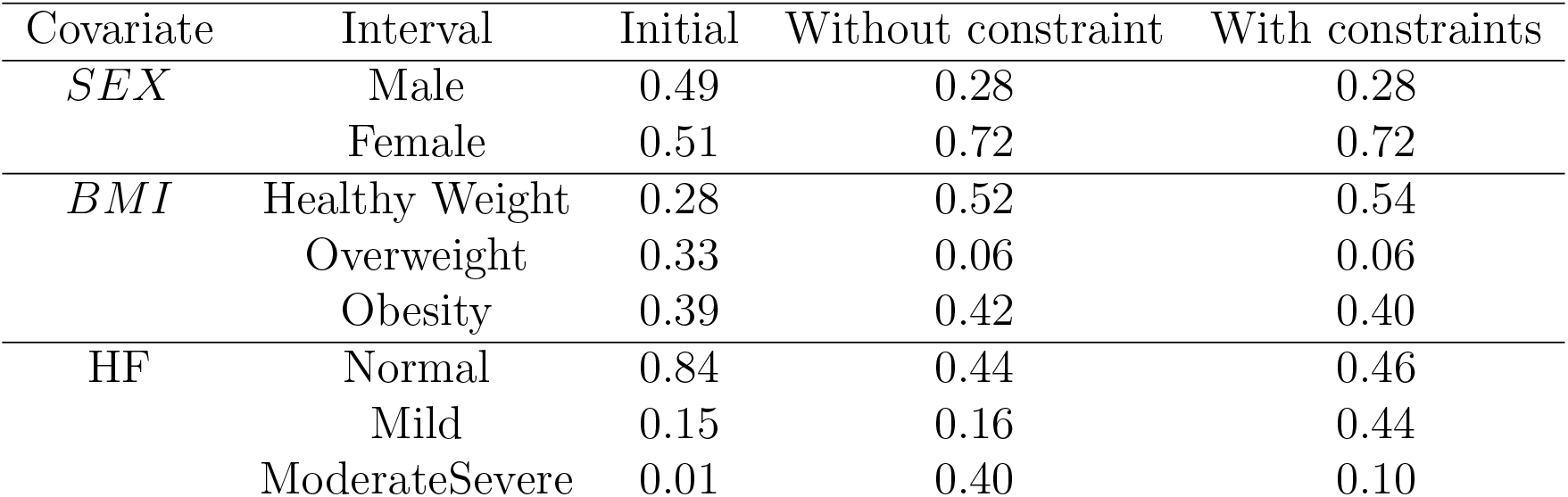
Hepatic case study - Marginal proportions for the covariate distributions for the D-criterion for the different sets of constraints.

